# A Cascaded Droplet Microfluidic Platform Enables High-throughput Single Cell Antibiotic Susceptibility Testing at Scale

**DOI:** 10.1101/2021.06.25.21259551

**Authors:** Pengfei Zhang, Aniruddha Kaushik, Kuangwen Hsieh, Sixuan Li, Shawna Lewis, Kathleen E. Mach, Joseph C. Liao, Karen C. Carroll, Tza-Huei Wang

## Abstract

Single-cell antibiotic susceptibility testing (AST) offers a promising technology by achieving unprecedented rapid testing time; however, its potential for clinical use is marred by its limited capacity for performing AST with scalable antibiotic numbers and concentrations. To lift the one antibiotic condition per device restriction common in single-cell AST, we develop a cascaded droplet microfluidic platform that uses an assembly line design to enable scalable single-cell AST. Such scalability is achieved by executing bacteria/antibiotic mixing, single-cell encapsulation, incubation, and detection in a streamlined workflow, facilitating susceptibility testing of each new antibiotic condition in 2 min after a 90 min setup time. As a demonstration, we test 3 clinical isolates and 8 positive urine specimens against 15 antibiotic conditions for generating antiprograms in ∼2 h and achieve 100% and 93.8% categorical agreement, respectively, compared to laboratory-based clinical microbiology reports which becomes available only after 48 h.

## 1. Introduction

The emergence of multi-drug resistant microorganisms is threatening to push humankind into the so-called “post-antibiotic era”^[1,2]^ and imperil human health worldwide. It has been projected that, without intervention, infectious diseases due to multi-drug resistant microorganisms could lead to ∼10 million annual global deaths by 2050,^[3]^ surpassing that of cancer. As the development of new antibiotics continues to stall,^[4]^ there is an urgent need for rapid identification and antibiotic susceptibility testing (AST) technologies that can promptly pinpoint the causative microorganisms and reveal their susceptibilities to antibiotics. Such diagnostic technologies are crucial for reducing empirical treatments for infectious diseases with broad-spectrum antibiotics, promoting evidence-based antibiotic prescriptions, enacting antibiotic stewardship, and facilitating resistance surveillance, thereby safeguarding existing antibiotics and minimizing the emergence of new multi-drug resistant microorganisms.^[5–12]^ To this end, significant advances in molecular diagnostics have created rapid identification technologies that are becoming turnkey in clinical microbiology. AST technologies, on the other hand, currently remain reliant on time-consuming culturing and thus still require about 48 h after sample collection.^[9]^ This means, even if the causative microorganisms can be rapidly identified, their AST results would remain unavailable, thus bottlenecking the complete diagnosis. This scenario underscores the critical but unmet need for developing rapid AST technologies that can work in sync with rapid identification technologies to mitigate the threat from multi-drug resistant microorganisms.

Among the technologies for advancing AST, microfluidics is particularly promising because miniaturization of AST in sub-microliter scale – orders of magnitude smaller than conventional AST – presents a powerful strategy for achieving rapid AST. Initial demonstrations of microfluidic AST that can be completed within a few hours,^[13–16]^ effectively illustrate the potential of this strategy. More recently, microfluidic single-cell technologies have pushed the boundaries of AST miniaturization to the scale of single bacteria. Through the confinement of single bacteria within nano-, pico-, or even femtoliter-scale channels, wells, or droplets, even a minute change in bacterial metabolism, genetic content, morphology or replication in response to antibiotic exposure can be detected. As a result, single-cell technologies have enabled some of the most rapid AST to date – some even in < 1 h.^[17–26]^ The reported assay turnaround times of these single-cell AST technologies approach the required time scale for preventing empirical prescription of broad-spectrum antibiotics. Unfortunately, most rapid single-cell AST technologies can only test one ^[18,20,22– 26]^ to four ^[19]^ antibiotic conditions per device. They are effective for rapidly calling the susceptibility or resistance for one antibiotic, but are unable to provide additional AST information beyond this binary call, which is a critical limitation.

The limited-antibiotic-condition testing capability afforded by rapid single-cell AST technologies deviates from the gold standard AST guidelines from the United States Clinical and Laboratory Standards Institute (CLSI) and European Committee on Antimicrobial Susceptibility Testing (EUCAST). Both guidelines recommend testing multiple antibiotics at predefined, clinically relevant titrations, producing an “antibiogram” with minimum inhibitory concentrations (MICs) range of the tested antibiotics against the causative microorganisms. The antibiogram is essential for clinical microbiology as it facilitates selection of antibiotics to ensure appropriate treatments and maintain antibiotic stewardship within healthcare settings while also enabling resistance surveillance by tracking pathogen MICs. Current rapid single-cell AST technologies lack the scalability to perform AST encompassing multiple antibiotics at multiple concentrations which severely limits their usefulness and translational potential for clinical microbiology. Thus, it is imperative to develop a rapid single-cell AST technology that possesses the ability to perform AST at scale as recommended by gold standard guidelines – and produce the essential antibiograms for clinical microbiology use.

In response, we have developed a microfluidic droplet-based platform – termed SCALe-AST (**<u>S</u>**ingle-**<u>C</u>**ell **<u>A</u>**ssembly **<u>L</u>**ine **AST**) – that achieves rapid and scalable single-cell

AST for multiple antibiotics at clinically-relevant titrations and produces an antibiogram with MICs (**Figure 1**). SCALe-AST employs a cascaded microfluidic design that integrates and simultaneously performs programmable bacteria/antibiotic mixing, single-cell encapsulation, incubation and detection in a streamlined and continuous process, akin to an assembly line. In doing so, SCALe-AST not only lifts the restriction on the number of antibiotic as well as concentrations processible by a single device, but also enables high-throughput single-cell AST at custom concentrations of antibiotics for generating complete antibiograms via a new statistics-based strategy. Moreover, a simple sample purification protocol can be appended to SCALe-AST for direct testing of clinical samples without conventional overnight, culture-based pathogen isolation. SCALe-AST could accurately measure the MICs for reference *E. coli* against common antibiotics and correctly assess the susceptibility or resistance for both reference strains and clinical isolates. Using SCALe-AST, we tested minimally processed urine specimens from patients with suspected urinary tract infections – one of the most prevalent infectious diseases^[27]^ that is often empirically treated with broad-spectrum antibiotics – and produced an antibiogram with MICs in ∼90 min (plus 2 min for each subsequent antibiotic condition) with 93.8% categorical agreement with the traditional antibiogram that would have required 48 h.

**Figure 1.**
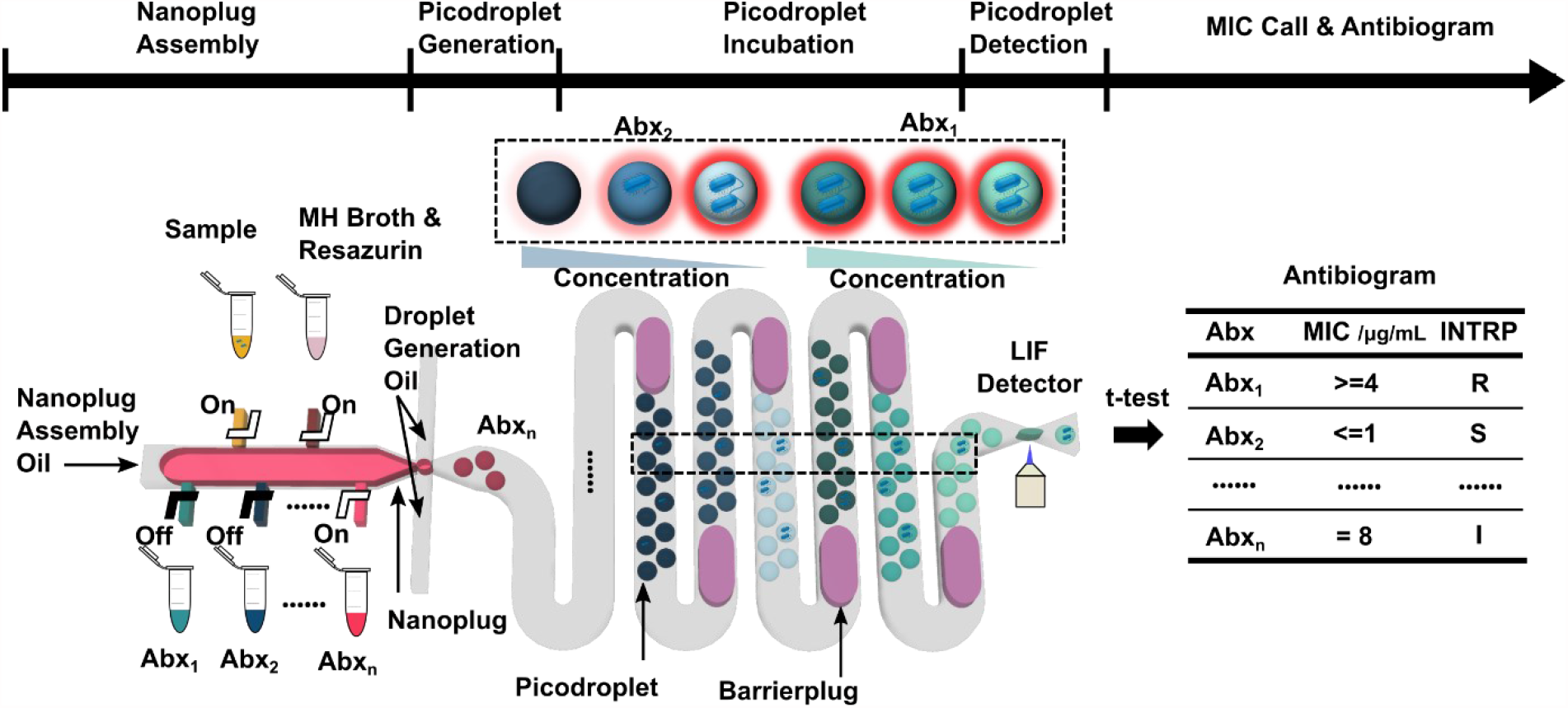
Overview of SCALe-AST. In SCALe-AST, an integrated device with programmable microvalves assembles bacteria sample, Mueller-Hinton II (MH) broth, resazurin, and antibiotics (denoted as Abx_1_, Abx_2_,…, Abx_n_) into nanoplugs and subsequently discretizes the nanoplugs into groups of picodroplets encapsulating single bacteria. Within the device, as each group of picodroplets flow through the built-in, 37 °C incubation channel, a barrierplug is introduced behind the picodroplets to keep them separated from adjacent groups of picodroplets, thus preventing cross-talk between different antibiotic conditions. The encapsulated single bacterium stops growing if it is susceptible to the applied antibiotic and the weakly fluorescent resazurin is reduced slowly, resulting in a weak fluorescence signal. In contrast, the bacterium proliferates if it is resistant to the applied antibiotic, and the weakly fluorescent resazurin is reduced to fluorescent resorufin quickly, resulting in a strong fluorescence signal within the picodroplet upon detection via a laser-induced-fluorescence (LIF) detector. By comparing picodroplet fluorescence intensity of different antibiotic concentrations, we are able to produce an antibiogram that provides the bacteria susceptibility categorization for multiple antibiotics with measured MICs.

## 2. Results

### 2.1. Design and workflow of SCALe-AST

At the heart of SCALe-AST is a fully integrated device (**Figure 2A** and **2B**). This device features a programmable microvalve-enabled nanoliter plug (*i*.*e*., nanoplug) assembler that combines samples, nutrient-rich Mueller-Hinton II broth (denoted as MH broth hereafter), a viability indicator (resazurin) and antibiotic into discrete nanoplugs at precisely controlled titrations (**Figure 1** and **2C(i)**). The assembled nanoplugs are discretized into picoliter droplets (*i*.*e*., picodroplets) through a flow-focusing junction (**Figure 1** and **2C(ii)**). SCALe-AST takes advantage of stochastic confinement of bacteria into picodroplets for single-cell AST via the detection of bacterial viability at various antibiotic concentrations. Rapid accumulation of metabolites from single cells within picodroplets facilitates assessment of bacterial viability at significantly shortened timescales as compared to conventional ensemble methods.^[23]^ Following each picodroplet group in our platform, we generate a blank nanoplug (denoted hereafter as barrierplug), which isolates and prevents cross-talk between adjacent droplet groups of different antibiotic conditions (**Figure 1** and **2C(iii)**). Each picodroplet group then flows through a 37 °C incubation region, before being individually detected using a laser-induced-fluorescence (LIF) system to characterize bacteria viability (**Figure 1** and **2C(iv)**). Following LIF detection, a database of all single-cell viability measurements is compiled for every antibiotic condition tested and statistically compared via t-tests to obtain MICs and construct clinically-useful antibiograms. The complete workflow is shown in **Video S1**.

**Figure 2.**
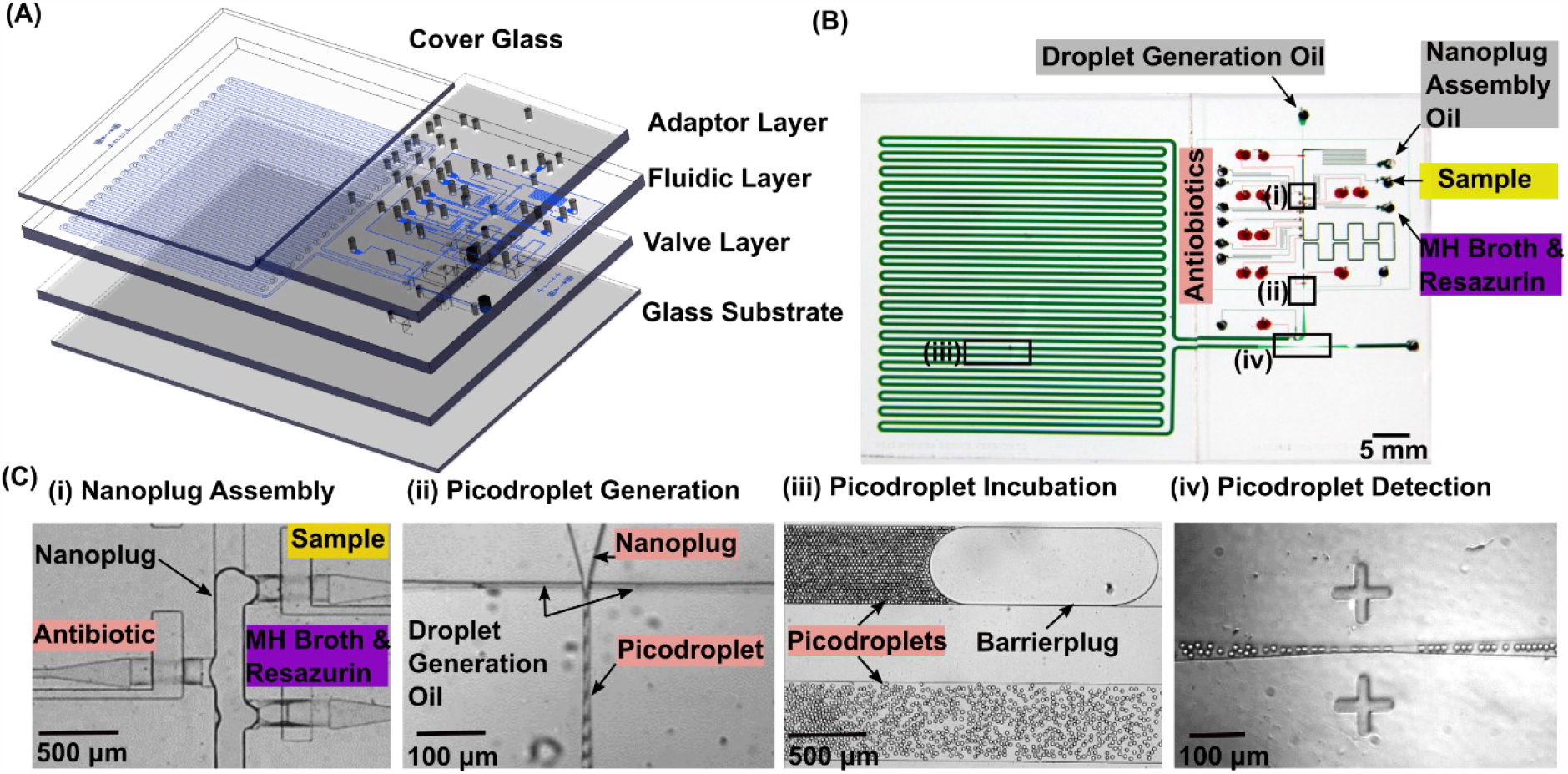
SCALe-AST device and its operation. (A) The device consists of a glass substrate, a valve layer, a fluidic layer, an adaptor layer, and a cover glass for preventing evaporation. (B) The device image shows the two key functional layers – the fluidic layer (green) and the valve layer (red) – and the inlets for injecting sample, reagents, and oils. (C) The device has integrated functionalities. (i) A nanoplug with a customized antibiotic/concentration is assembled by microvalve actuation. (ii) The assembled nanoplug is discretized into picodroplets using a flow-focusing junction. (iii) In-line incubation is integrated to streamline droplet manipulation with a barrierplug introduced for droplet group separation. (iv) Picodroplets flow through a constricted detection channel, where they are detected using a laser-induced-fluorescence (LIF) detector (not shown).

### 2.2. Device optimization for nanoplug assembly and picodroplet generation

Reliable performance of SCALe-AST requires implementation of salient device designs to ensure rapid but thorough mixing of reagents within nanoplugs, stable generation of monodisperse picodroplets of appropriate volume and precise and reproducible reagent concentrations in the end. First, to accelerate mixing within the assembled nanoplug and speed up multiplex AST, we determined the maximum mixing channel height to minimize the nanoplug length, which is the rate limiting factor for mixing (**Figure S1A**). With the channel height of ∼120 µm, the nanoplug mixing can be completed in 5 s – confirmed by picodroplets with consistent contents (CV=5.70%) – as opposed to ∼50 s mixing in a mixing channel with a height of ∼60 µm (**Figure S1B** and **S1C**). Next, we optimized the flow-focusing junction in order to achieve stable generation of monodisperse picodroplets of appropriate volume. We recognized that reducing the picodroplet volume would increase the relative concentration of single-cell metabolic product and thus accelerate our measurements of single-cell viability.

Therefore, we sought to minimize picodroplet size by decreasing the height of the flow-focusing junction (**Figure 2C(ii)**). We discovered that for channel heights less than 22 µm, the ceiling of the microchannel could easily collapse into the floor, hindering stable droplet generation. We therefore settled on a 22-µm-tall and 10-µm-wide flow-focusing junction to generate monodisperse 8-pL picodroplets (CV = 3.6%) (**Figure S2A** and **S2B**). Of note, we coated the walls of our channels with a hydrophobic Rain-X layer, which we had empirically found to significantly improve the stability of picodroplet generation (**Figure S3** and **Video S2**). With additional device optimizations (see S1 in Supporting Information), we finally demonstrated that concentrations can be generated in picodroplets with high precision and reproducibility by using our device. We proved this point by generating multiple picodroplet groups containing linearly increasing concentrations of resazurin and measuring the corresponding fluorescence signals from these picodroplets with high linearity (**Figure S4**).

### 2.3. Barrierplugs for spatial separation between picodroplet groups

As a group of picodroplets freshly discretized from a nanoplug flow through the incubation channel under pressure-driven flow, the picodroplets experience different flow velocities across the width of the channel and as a result disperse along the channel, a phenomenon commonly referred to as Taylor dispersion.^[28]^ In the absence of a separating mechanism, multiple groups of picodroplets – all experiencing Taylor dispersion – would mix (**Figure 3A(i)** and **Video S3**) and lead to cross-talks in fluorescence detection of picodroplets (**Figure 3A(ii)**). Such cross-talks would severely complicate SCALe-AST because the fluorescence intensity alone in each picodroplet cannot simultaneously reveal the presence of bacteria, the concentration of the antibiotic, and the response of the bacteria to the antibiotic concentration. In theory, fluorescence coding (*i*.*e*., assigning a distinct concentration of a second fluorophore to each group of picodroplets containing the same antibiotic concentration) can alleviate cross-talks in fluorescence detection, but significant picodroplet dispersion still means widely varied incubation time for the bacteria exposed to an antibiotic concentration, and thus prevents standardized single-cell AST. Finally, lengthening the time gap between adjacent picodroplet groups may reduce cross-talks, but the challenge of widely varied incubation time would persist while lengthy downtimes would significantly increase the assay time and severely hamper the usefulness of SCALe-AST.

**Figure 3.**
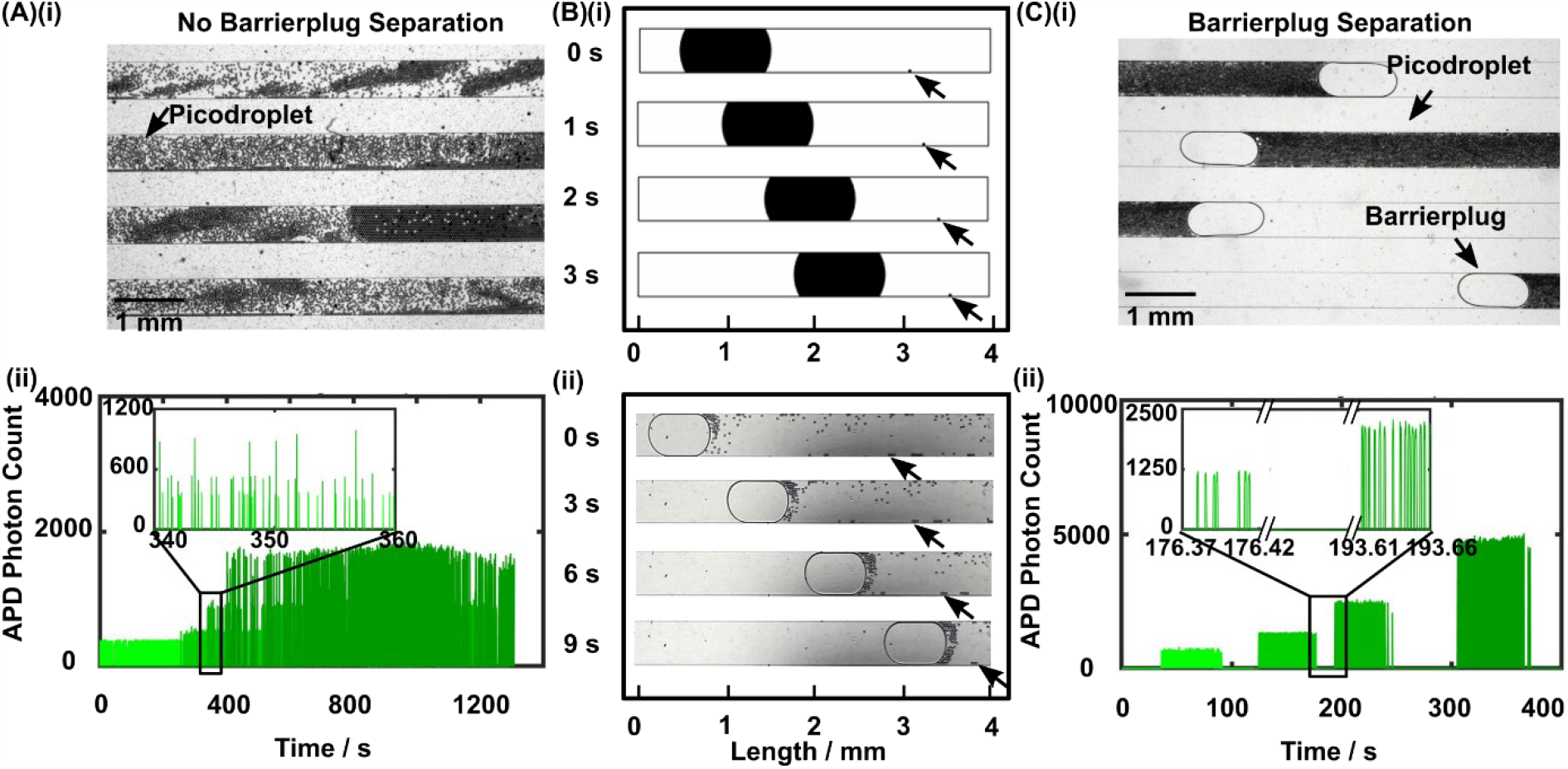
Barrierplugs for separation of picodroplet groups. (A)(i) In the absence of barrierplugs, picodroplets can disperse as they flow through the incubation channel and cause significant mixing among picodroplet groups. (ii) Such dispersion and mixing can be fluorescently detected as picodroplet groups with distinct fluorophore concentrations would yield a fluorescence trace showing mixed peaks (*i*.*e*., picodroplets) with distinct avalanche photodiode (APD) photon counts (*i*.*e*., fluorescence intensities). (B)(i) Two-dimensional water-oil multiphase flow CFD simulation reveals that, within an oil-filled, 500-µm-wide microchannel (white) and under pressure-driven flow, a water barrierplug (black ellipse, 1000 μm × 500 μm semiaxes) that seals the entire width of the channel flows faster than a downstream water picodroplet (black circle, 25 μm in diameter, indicated by black arrows) that is positioned at the wall of the channel, as indicated by the decreasing distance between them over time (t = 0, 1, 2, and 3 s). (ii) Experimental observation within the incubation channel of a SCALe-AST device corroborates with the simulation, as a barrierplug indeed catch up to picodroplets at the channel wall over time (t = 0, 3, 6, and 9 s). (C)(i) In the presence of barrierplugs, which prevent dispersion of picodroplets, picodroplet groups therefore become tightly packed and well separated from each other. (ii) Well separated picodroplet groups due to the addition of barrierplugs can be fluorescently detected as picodroplet groups with distinct fluorophore concentrations could now yield a fluorescence trace showing well separated groups of peaks with distinct photon counts.

As a solution to Taylor dispersion of picodroplets and the resulting complications, we hypothesized that the addition of a nanoplug would mitigate Taylor dispersion and provide a separating barrier between adjacent picodroplet groups. To test this hypothesis, we first created a two-dimensional multiphase computational fluid dynamics (*i*.*e*., CFD) simulation using COMSOL. In our model, we placed a picodroplet downstream to an elliptical “barrierplug” that seals the entire width of the channel. The picodroplet was positioned at the edge of the channel to simulate the lagging picodroplets with slowest flow velocity in a pressure-drive flow. Upon initiating pressure-driven flow, the barrierplug proceeds with a much higher velocity than the picodroplet. As a result, the faster barrierplug is able to gradually catch up with the lagging picodroplet (**Figure 3B(i)** and **Video S4**). Our simulation therefore supports the hypothesis that barrierplugs could be employed to push trailing picodroplets forward, counteract Taylor dispersion, and provide a barrier between adjacent groups of picodroplets.

We then experimentally verified the function of barrierplugs in our device. We added a microvalve-controlled inlet downstream to the flow-focusing junction (**Figure S5**) to inject a barrierplug containing only phosphate-buffered saline (PBS) behind every group of picodroplets. Consistent with our model, barrierplugs indeed flowed faster than picodroplets, especially those close to the sidewalls of the channel (**Figure 3B(ii)**, and **Video S1**). As a result, as the barrierplug flowed through the incubation channel, it would push trailing picodroplets forward until a tightly packed group of picodroplets was formed (**Figure 3C(i)**). Moreover, because the picodroplets were confined between barrierplugs, we could minimize cross-talk between groups of picodroplets that contained different concentrations of reagents. We confirmed picodroplet grouping by observing clearly separated clusters of fluorescence peaks with increasing intensities that originate from distinct groups of picodroplets containing increasing concentrations of fluorescein (**Figure 3C(ii)**). The tight packing enabled by barrierplugs ensured nearly identical droplet incubation duration with minimal variation, thus facilitating standardized single-cell AST (**Figure S6**, see S2 in Supporting Information for details). This enabling feature ensured that we could assemble and process unique groups of picodroplets in continuous flow and as a result drastically shorten the downtime for screening multiple antibiotic conditions in AST (see calculation in S3 of Supporting Information).

### 2.4. Single-cell AST and statistics-based MIC measurements

Following device optimization, we used the CLSI-recommended reference strain of *E. coli* (ATCC 25922) and trimethoprim-sulfamethoxazole (a first-line antibiotic; hereafter abbreviated as SXT based on the American Society for Microbiology convention) to establish single-cell AST in our device. The CLSI MIC for SXT against *E. coli* ATCC 25922 is < 0.5 µg mL^-1^ trimethoprim and 9.5 µg mL^-1^ sulfamethoxazole (conventionally referred to as 0.5/9.5 µg mL^-1^). We therefore tested around this MIC with the CLSI convention of a 2-fold titration series (*i*.*e*., 0.12/2.4, 0.25/4.8, 0.5/9.5, 1/19, and 2/38 µg mL^-1^). We also included a no-SXT control to serve as the reference in our single-cell AST. In the SCALe-AST device, we sequentially assembled 6 ∼80 nL nanoplugs that contained ∼10^7^ CFU mL^-1^ *E. coli*, MH broth, 100 µM resazurin, and the SXT titers in increasing order. The 6 nanoplugs were then sequentially discretized into 6 groups of ∼10000 8-pL picodroplets separated by barrierplugs. The picodroplets then flowed for 90 min through the incubation channel that posited on a custom 37 °C heat block. Finally, the picodroplets flowed one by one through the detection channel, where the fluorescence intensity from each picodroplet was measured by our LIF detector, eventually resulting in a trace of fluorescence intensity peaks from 6 distinct groups of picodroplets (**Figure 4A**). From the trace, we could distinguish high-intensity (*i*.*e*., positive) picodroplets containing strongly fluorescent resorufin that was reduced from weakly fluorescent resazurin by viable *E. coli* (**Figure S7**) from the low-intensity (*i*.*e*., negative) picodroplets containing only weakly fluorescent resazurin due to either no *E. coli* or SXT-susceptible *E. coli* (**Figure 4A, insets**).

**Figure 4.**
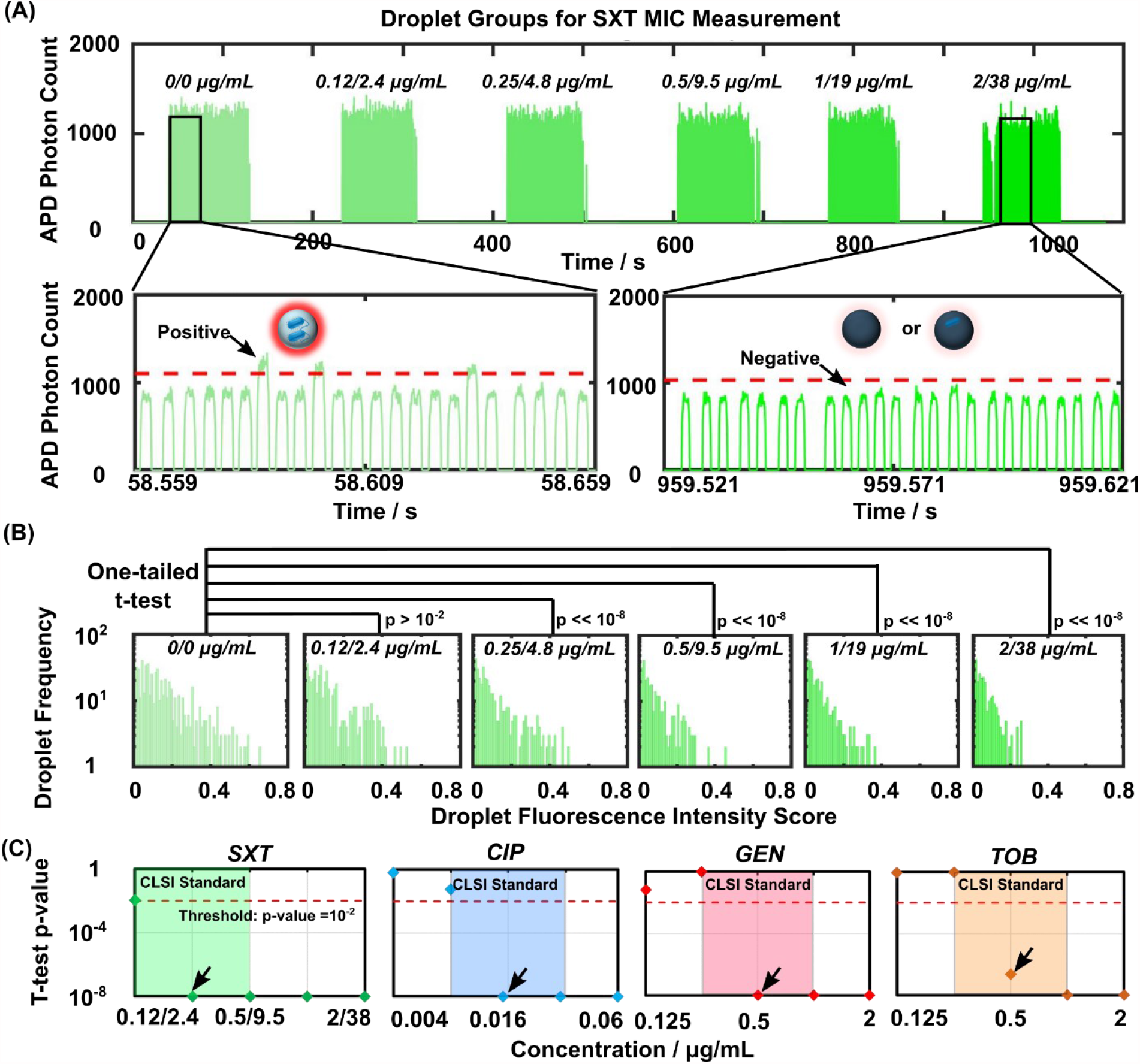
Single-cell AST and statistics-based minimum inhibitory concentration (MIC) measurements. (A) Using reference strain of *E. coli* (ATCC 25922) and titrations of trimethoprim-sulfamethoxazole (SXT; a first-line antibiotic) defined by the Clinical Laboratory Standards Institute (CLSI), SCALe-AST produces a fluorescence trace that shows distinct picodroplet groups corresponding to the SXT concentrations, which is used for MIC measurement. Zoom in: picodroplets above threshold are defined as ‘positive’, where the resazurin is reduced to highly fluorescent resorufin by *E. coli*, while droplets below threshold are defined as ‘negative’ where the resazurin remain weakly fluorescent due to either no *E. coli* or SXT-susceptible *E. coli* killed by the antibiotic. (B) The fluorescence intensity of each positive picodroplet from each group is converted a fluorescence intensity score (see Methods). As the SXT concentration increases, the positive picodroplet population becomes smaller and the fluorescence intensity scores become lower. Between the no-SXT control and each SXT concentration, these differences are assessed via one-tailed Student’s t-test, which provides unique strategy for measuring the MIC. For the t-tests, N=584, 474, 434, 330, 347 and 293 for 0/0, 0.12/2.4, 0.25/4.8, 0.5/9.5, 1/19, 2/38 µg/mL, respectively. (C) Here, the MICs (indicated by black arrows) are defined as the lowest antibiotic concentration that differs significantly (p << 10^−2^) from the corresponding no-antibiotic control. In this and subsequent figure, p-values ≤ 10^−8^ are uniformly displayed at 10^−8^ for clarity. The MICs for SXT, CIP, GEN, and TOB against *E. coli* (ATCC 25922) measured via SCALe-AST are 0.25/4.8 µg mL^-1^, 0.015 µg mL^-1^, 0.5 µg mL^-1^, and 0.5 µg mL^-1^, respectively, which match with standard MICs. Results presented in this and subsequent figures can be found in Data S1 in supplementary materials.

As SCALe-AST yielded ∼10000 fluorescence intensity peaks per condition, we devised a novel analysis strategy to fully leverage the data points towards measuring the MIC. This strategy contrasts from previous droplet-based AST, which presented AST results with a single value such as the percentage of positive picodroplets even if thousands of single-cell-containing droplets were detected. Specifically, in our strategy, we first calculated the fluorescence intensity score of each positive picodroplet from its measured fluorescence intensity (see Methods for details). We used histogram analysis to visualize that as the concentration of SXT increased, both the number and the intensity score of positive picodroplets decreased (**Figure 4B**) – a reflection of increasingly inhibited *E. coli* growth due to increasing SXT concentrations. We then performed one-tailed Student’s t-test to quantitatively compare the intensity scores from the no-SXT control and each of the 5 SXT concentrations and subsequently employed the resulting p-values as the basis for measuring the MIC. At 0.12/2.4 µg mL^-1^ SXT, we found a lack of significant difference (p > 10^−2^ or 0.01 a common benchmark p-value) from the no-SXT control, which corresponds to *E. coli* growth at this concentration. Conversely, at SXT concentrations ≥ 0.25/4.8 µg mL^-1^, we found highly significant differences (p << 10^−8^) from the no-SXT control that are indicative of significantly inhibited *E. coli* growth (**Figure 4B**) and are consistent with the CLSI MIC. This agreement thus provides initial support for our novel MIC measurement strategy of leveraging t-tests to analyze single-cell AST results acquired via SCALe-AST.

To further verify SCALe-AST and demonstrate its scalability, we measured the MICs of SXT and 3 common antibiotics – ciprofloxacin (CIP), gentamicin (GEN), and tobramycin (TOB) – against the ATCC reference *E. coli* within a single device. The CLSI MICs for CIP, GEN, and TOB, conventionally shown in 2-fold titrations, are 0.008 to 0.03 µg mL^-1^, 0.25 to 1 µg mL^-1^, and 0.25 to 1 µg mL^-1^, respectively. For ensuring accurate MIC measurements in our device, we tested the aforementioned SXT concentrations, and we added a 2-fold higher concentration and a 2-fold lower concentration around the CLSI MICs and tested CIP from 0.004 to 0.06 µg mL^-1^, GEN from 0.12 to 2 µg mL^-1^, and TOB from 0.12 to 2 µg mL^-1^. Here, for each antibiotic, we also added a group of “spatial coding picodroplets” (for indicating the beginning of a new antibiotic in our fluorescence intensity trace) and two groups of no-antibiotic control picodroplets, resulting in 32 consecutive groups of picodroplets within the device (**Figure S8**). Importantly, the continuous-flow, assembly-line operation of our device maintained fast turnaround. While the first condition was tested in 90 min, each subsequent condition only added ∼2 min to the turnaround time, for a turnaround time of ∼150 min (**Figure S8**). After calculating intensity scores and performing t-tests, our SCALe-AST MIC curves based on t-test p-values resembled conventional MIC curves (**Figure 4C**). In particular, SXT ≥ 0.25/4.8 µg mL^-1^, CIP ≥ 0.015 µg mL^-1^, GEN ≥ 0.5 µg mL^-1^, and TOB ≥ 0.5 µg mL^-1^ were significantly different from their respective no-antibiotic controls (p << 10^−2^, **Figure 4C**, threshold at p = 10^−2^ indicated by red dotted line). Our measured MICs for SXT, CIP, GEN, and TOB were therefore 0.25/4.8 µg mL^-1^, 0.015 µg mL^-1^, 0.5 µg mL^-1^, and 0.5 µg mL^-1^, respectively (**Figure 4C**, black arrows) – in full agreement with CLSI MICs for this *E. coli* reference strain (**Figure 4C**, shaded regions). Replicating single-cell AST in 2 more devices, our measured MICs remained congruent with CLSI MICs (**Figure S9**). These results demonstrate that single-cell AST and t-test-based analysis in SCALe-AST can accurately and reproducibly measure the MICs of the ATCC reference *E. coli* against common antibiotics, which present important benchmarks for developing new AST technologies.

### 2.5. Antibiotic susceptibility categorization for clinical isolates

We next moved on to demonstrating the applicability of SCALe-AST in categorizing the antibiotic susceptibilities of clinical isolates (**Figure 5A**). Here, following the CLSI guideline for testing clinical isolates, AST was performed by challenging the bacteria at two (e.g., SXT) or three (e.g., CIP, GEN, TOB) interpretive breakpoint concentrations defined by the CLSI to categorize the bacteria as either susceptible (S) or resistant (R) for SXT, or as S, intermediate (I), or R for CIP, GEN, and TOB. Performing AST at these interpretive breakpoint concentrations produces results in the following three categories: S when the measured MIC is less than or equal to the lowest interpretive breakpoint concentration, I when the measured MIC is equal to the middle concentration, or R when the measured MIC is greater than or equal to the highest concentration. These categories provide the clinically relevant information that physicians need to administer treatments.^[29]^ As such, even in the absence of precise values, the MICs measured from AST at interpretive breakpoint concentrations are clinically relevant (referred to as “clinical MICs” hereafter for differentiating from the previous MICs of the ATCC reference *E. coli*).

**Figure 5.**
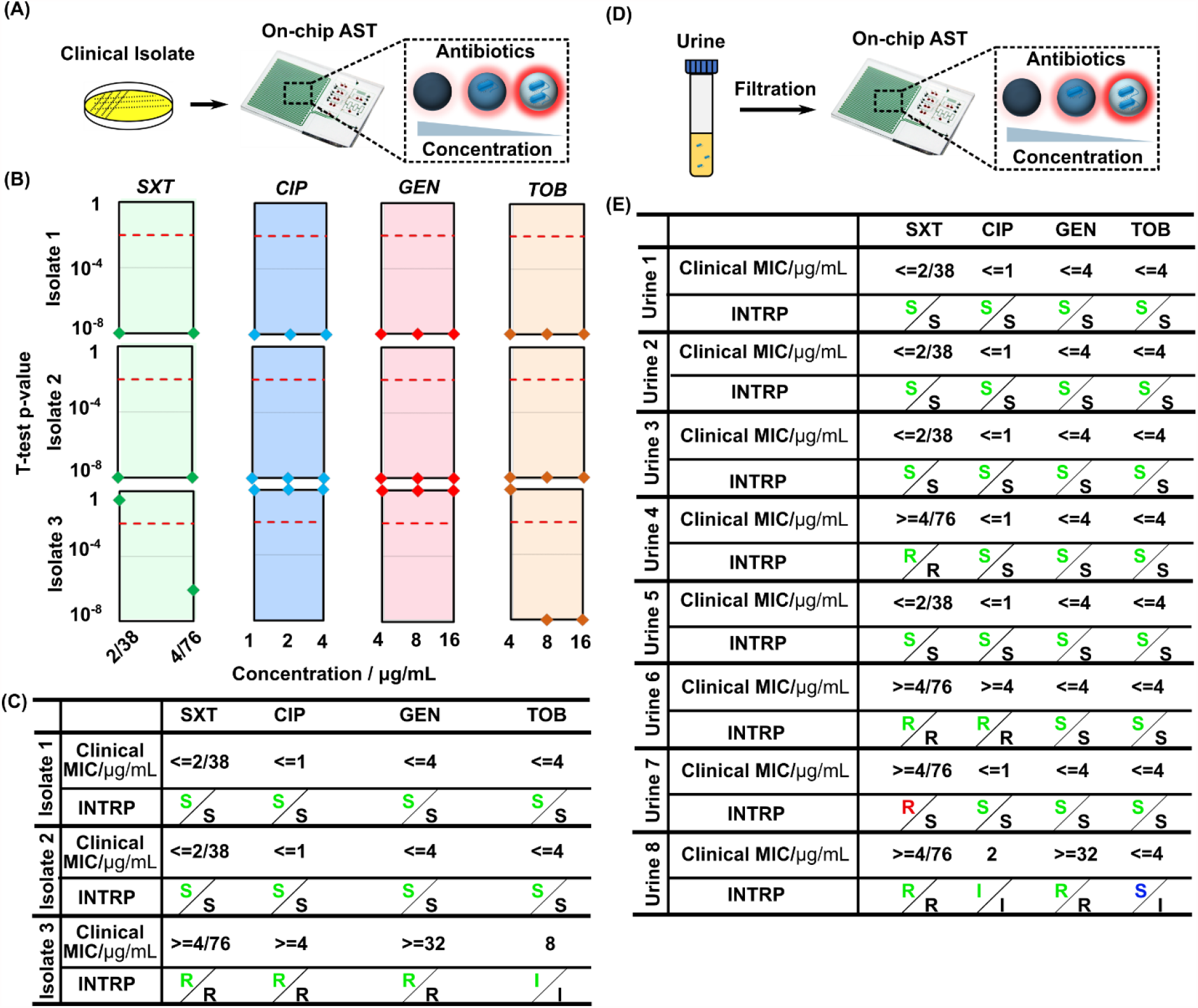
Evaluation of SCALe-AST with clinical isolates and clinical urine samples. (A) SCALe-AST is first evaluated with clinical isolates at the CLSI-defined interpretive breakpoint concentrations for each antibiotic. (B) For 3 clinically isolated *E. coli* against SXT, CIP, GEN, and TOB, SCALe-AST produces t-test-based results, which are used to (C) measure clinical MICs and categorize susceptibilities. The SCALe-AST antibiogram (green “susceptible/intermediate/resistant”, or S/I/R) matches clinical report (black S/I/R) with 100% concordance. (D) SCALe-AST is coupled to a simple filtration protocol and evaluated with clinical urine specimens without prior clinical isolation, again at the CLSI-defined interpretive breakpoint concentrations for each antibiotic. (E) For 8 clinical urine specimens against SXT, CIP, GEN, and TOB, SCALe-AST (green S/I/R) achieves 93.8% concordance with the clinical report (black S/I/R), and 1 minor error (susceptible call for intermediate category; blue), 1 major error (resistant call for susceptible phenotype; red), and 0 very major error (susceptible call for resistant phenotype).

Prior to testing clinical isolates, we established the workflow for performing interpretive breakpoint testing for SXT, CIP, GEN, and TOB in our device with the ATCC reference *E. coli* and a ATCC multidrug-resistant *E. coli* strain. For testing both *E. coli* strains, we followed the CLSI-recommended interpretive breakpoint concentrations for *Enterobacterales* (the taxonomic order to which *E. coli* belongs): 2/38 and 4/76 µg mL^-1^ for SXT, 1, 2, and 4 µg mL^-1^ for CIP, 4, 8, and 16 µg mL^-1^ for GEN, and 4, 8, and 16 µg mL^-1^ for TOB. Each strain was tested against 23 consecutive groups of picodroplets that include antibiotic titers, no antibiotic controls, and spatial codes in a single device and the turnaround time was ∼2 h. After single-cell AST and t-test-based analysis, SCALe-AST correctly identified the ATCC reference *E. coli* as S to the four antibiotics and the ATCC multidrug-resistant *E. coli* as R to the four antibiotics (**Figure S10**). Moreover, we triplicated interpretive breakpoint testing in independent devices for both reference *E. coli* and multi-drug resistant *E. coli* and consistently called the correct susceptibility categories for the four antibiotics for both *E. coli* strains (**Figure S10**), thus demonstrating robust and reproducible interpretive breakpoint testing in SCALe-AST.

We tested 3 de-identified *E. coli* clinical isolates provided by the Johns Hopkins Hospital (JHH) Clinical Microbiology Laboratory (IRB00189525), each against SXT, CIP, GEN, and TOB at the CLSI-recommended interpretive breakpoint concentrations for *Enterobacterales* in a SCALe-AST device. For isolates 1 and 2, the t-test p-values from all antibiotic concentrations – even the lowest concentrations corresponding to the susceptible breakpoints – were << 10^−2^ (**Figure 5B**). Accordingly, the measured clinical MICs were ≤ 2/38 µg mL^-1^ for SXT, ≤ 1 µg mL^-1^ for CIP, ≤ 4 µg mL^-1^ for GEN, and ≤ 4 µg mL^-1^ for TOB, which corresponded to S for all four antibiotics for both isolates (**Figure 5C**). In contrast, for isolate 3, the t-test p-values were << 10^−2^ only for 4/76 µg mL^-1^ SXT and 8 and 16 µg mL^-1^ TOB (**Figure 5B**). SCALe-AST thus measured the clinical MICs as ≥ 4/76 µg mL^-1^ for SXT, ≥ 4 µg mL^-1^ for CIP, ≥ 16 µg mL^-1^ GEN, and 8 µg mL^-1^ TOB, which corresponded to R for SXT, CIP, and GEN, and I for TOB for this isolate (**Figure 5C**). Compared to clinical antibiograms provided by JHH, SCALe-AST achieved 12 out of 12 categorical agreements (**Figure 5C**). These results not only highlight SCALe-AST’s performance for testing clinical isolates, but also demonstrate an advantage of SCALe-AST over common single-cell AST platforms that only test a single antibiotic at a single concentration, which would be unable to make the intermediate categorization or measure clinical MICs for multiple antibiotics.

### 2.6. Clinical isolation-free SCALe-AST from urine samples

For the final demonstration of SCALe-AST, we focused on testing clinical urine samples because urinary tract infections (UTIs) are among the most prevalent infectious diseases and demand advances in diagnostic solutions. Indeed, ∼50% women suffer from UTIs at least once during their lifetime.^[30]^ Meanwhile, physicians are often forced to initiate empirical treatments for UTIs with broad-spectrum antibiotics before the antibiogram becomes available after 48 – 72 hours – a practice that can worsen patient outcomes, drive up healthcare costs, and fuel the emergence of multi-drug-resistant pathogens.^[1,31]^ We were thus motivated to shorten the total turnaround time of our SCALe-AST by obviating clinical isolation. Meanwhile, we recognized that transferring bacteria from urine into MH broth is critical because non-bacterial impurities (e.g., epithelial cells, white blood cells, proteins, and metabolites) and chemical additives (e.g., sodium borate) in clinical urine samples would interfere with our resazurin-based AST. As a solution for both demands, we devised a facile filtration protocol in which we first filtered the urine sample through a gradient-based syringe filter (iPOC^DX^ Primecare, 35 to 5 μm) to remove impurities larger than bacteria, then passed the filtrate through a 0.22-µm-pore syringe filter to trap bacteria and discard urine, and finally injected MH broth in the reverse direction through the syringe filter to recover bacteria in MH broth (**Figure S11**, see Methods for details). This protocol, which allowed us to recover up to 66% spiked *E. coli* from urine into MH broth within 10 min,^[32]^ paved the way for us to demonstrate the feasibility of clinical isolation-free SCALe-AST.

We finally used clinical isolation-free SCALe-AST to test 8 de-identified UTI-positive urine samples provided by the JHH Clinical Microbiology Laboratory (IRB00189525). Urine specimens that represented uncomplicated UTIs were selected for our study. Each urine specimen was subject to the clinical standard diagnostic workflow that included clinical isolation via plating and AST via the BD Phoenix instrument which uses broth microdilution testing. Clinical AST reports for each specimen required at least 48 h from patient collection. SCALe-AST was used to test refrigerated urine specimens that had freshly entered the clinical workflow, in order to obtain comparable results within a small fraction of the time using our filtration protocol and SCALe-AST (**Figure 5D**). Upon comparison with the clinical antibiograms, we achieved 30 out of 32, or 93.8%, categorical agreement with only 1 minor error (*i*.*e*., S by SCALe-AST but I in clinical antibiogram), 1 major error (*i*.*e*., R by SCALe-AST but S in clinical antibiogram), and 0 very major error (*i*.*e*., S by SCALe-AST but R in clinical antibiogram) (**Figure 5E**). These results provide further support that SCALe-AST can reliably perform interpretive breakpoint testing and accurately categorize antibiotic susceptibilities. Furthermore, these results also show the feasibility of using filtration and SCALe-AST to directly analyze clinical urine specimens for uncomplicated UTIs without time-consuming clinical isolation.

## 3. Conclusion

SCALe-AST is the first droplet microfluidic platform that performs rapid, single-cell AST within picoliter-scale droplets at the clinically appropriate scale of antibiotic concentrations in an assembly-line-like workflow to generate antibiograms and MICs. The development of SCALe-AST incorporates three enabling concepts. First, the cascaded design of the microfluidic device allows on-demand nanoplug assembly, picodroplet generation, picodroplet incubation, and picodroplet detection to simultaneously take place akin to an assembly line, thus enabling scalable single-cell AST within a single device. This design lifts the constraint on the number of antibiotics or concentrations that can be tested in a single device, which facilitates MIC measurements and antibiotic susceptibility categorization. Second, with both computational simulation and experiment results, we demonstrated our barrierplug design contained picodroplet dispersion and achieved clear separation of different picodroplet groups. As a result, barrierplugs facilitates the immediate setup of a new picodroplet group as the preceding droplet groups undergo incubation and detection without concern of cross-talk, thus minimizing the downtime in SCALe-AST. Third, t-test-based analysis in SCALe-AST enhances already powerful single-cell AST. Single-cell AST is inherently rapid because the confinement of single bacteria in picodroplets allows rapid accumulation of metabolic products and therefore shortens the assay time. SCALe-AST further leverages high-throughput and quantitative detection of ∼10000 picodroplets from each antibiotic concentration and unlocks t-test-based analysis allowing detection of minute changes of *E. coli* growth across different antibiotic concentrations and the rapid time. Successful implementation of these concepts allows a single device to process and analyze up to 32 groups of ∼10000 picodroplets, while maintaining a fast susceptibility testing of each new antibiotic condition in 2 min after a 90 min setup time.

In our proof-of-concept demonstration, we used SCALe-AST to test urine samples containing *E. coli*, without pathogen isolation, against four common antibiotics and report the antibiograms and the clinical MICs. We focused on UTIs since 65-75% of cases are caused by only *E. coli*.^[27]^ Even without pathogen identification capabilities, SCALe-AST’s capacity for direct, isolation-free, rapid AST can provide actionable diagnostic results for a significant portion of UTIs. We also note that the four antibiotics we tested were supported by a clinically-oriented rationale. Currently, SXT and CIP are among the most frequently prescribed antibiotics in the outpatient setting for UTIs.^[33]^ Unfortunately, the overuse of SXT and CIP has caused continuously increased resistance level among uropathogens.^[33,34]^ As an alternative for SXT and CIP, single-dose aminoglycoside (including GEN, TOB) therapy has been demonstrated as an adequate treatment for UTI.^[35,36]^ By generating a rapid antibiogram for these four antibiotics using our platform, we not only provide a more complete susceptibility spectrum for treatment guidance, but also avoid SXT and CIP overuse for better antibiotic stewardship.

SCALe-AST offers a promising new path for elevating the throughput of single-cell-based testing and performing such testing at the scale required for diagnostic testing for infectious diseases. With additional developments, we envision that SCALe-AST may inform clinical-decision-making for other infectious diseases. SCALe-AST could work in tandem with rapid pathogen identification platforms to achieve pathogen identification and AST simultaneously given the fast development and wide availability of those platforms.^[9]^ Moreover, SCALe-AST can further minimize the category error for clinical specimen testing with alternative bacteria recovery and enrichment methods to concentrate bacteria.^[32]^ This is especially important because clinical samples have unknown bacterial loads. With more extensive follow-up investigation, SCALe-AST has potential to guide evidence-based treatment, reduce empirical antibiotic prescription and enable better antibiotic stewardship in the future.

## 4. Materials and methods

### Master Mold Fabrication

SCALe-AST consists of two key functional layers – a fluidic layer for nanoplug assembly, picodroplet generation, incubation and detection, and a valve layer for controlling the creation and movement of these nanoplugs and picodroplets. Masks for the fluidic and valve control layers were designed in AutoCAD (Autodesk Inc., San Rafael, CA, USA). Mask patterns were printed onto high-quality transparencies at 20,000 dpi by CAD/Art Services, Inc. (Bandon, OR, USA).

Master molds for both fluidic and valve control layers of the device were microfabricated via standard photolithography on 4 in. silicon wafers (Polishing Corporation of America, Santa Clara, CA, USA). To construct the mold for the fluidic channel, we firstly fabricate the segments of channel that intersects the polydimethylsiloxane (PDMS) microvalves. Specifically, SPR-220-7 (positive photoresist; Microchem Corp., Newton, MA, USA) was spin-coated to a height of ∼24 µm and then melted at 200 °C to form a rounded cross-section, which allows the microfluidic channels to be effectively closed off by the PDMS membrane upon microvalve closure. For fabricating the remaining fluidic channels that do not intersect PDMS microvalves, we designed different channel heights for different purposes, including nanoplug assembly and mixing (∼120 µm), picodroplet generation and detection (∼22 µm) and picodroplet incubation ((∼60 µm)). All these layers were fabricated using SU8-3025 (Microchem Corp., Newton, MA, USA) photoresist and they were stacked, aligned and patterned via photolithography using common alignment marks on the wafer. For each layer, every step required for photolithography (including soft bake, UV exposure and post exposure bake) was conducted except development, which was performed after the last layer was patterned. This strategy was effective to avoid air bubble generation caused by preexisting developed features. For the valve control mold, a single layer of SU8-3025 was spun to a height of ∼30 µm and patterned via photolithography.

### Microfluidic Device Fabrication

SCALe-AST devices were fabricated in a similar way to our previous protocol^[37]^ (**Figure S12**). Briefly, a thin valve control layer (∼50 μm) was fabricated by spin-coating 15:1 PDMS (SYLGARD 184 Silicone Elastomer Kit, Dow Corning, Midland, MI, USA) onto the valve control mold at 1000 rpm for 60 s and curing for 12 min. at 80 °C. A thicker (∼ 1 mm) sacrificial PDMS layer was formed by spin-coating 6:1 PDMS on a blank wafer at 100 rpm for 60 s and baked at 80 °C for 7 min. This PDMS sacrificial layer was reversibly bonded on top of the thin PDMS valve control layer via baking for 8 min. at 80 °C and served as a structural support to peel the valve control layer from its mold. Then, the valve control layer was permanently bonded to a cover glass (75 mm in length, 50 mm in width, and 130 μm in thickness, Ted Pella, Redding, CA) using oxygen plasma treatment (42 W, 500 mTorr, 45 s). Then the sacrificial PDMS layer could be easily peeled off and discarded. At the same time, a ∼1 mm thick fluidic layer was fabricated by spin-coating 10:1 (base to curing agent ratio) PDMS onto the fluidic mold at 100 rpm for 60 s and curing for 15 min. at 80 °C. The fluidic layer was then permanently bonded to the valve control layer via oxygen plasma treatment after manually aligning using a stereoscope. For easy macro-to-micro interface, a 5-mm thick PDMS layer with inlet and outlet accesses (*i*.*e*., adaptor layer) were fabricated and bonded with the microvalve control part of fluidic layer while the incubation channel part of fluidic layer was sealed with a cover glass to minimize droplet evaporation during incubation (**Figure 2, Figure S12**, and **S13**). The completed device was baked for at least 24 hours at 80 °C. In order to ensure optimal hydrophobicity to minimize liquid sticking and stabilize picodroplet generation, a commercial hydrophobic coating agent (Rain-X® Original, ITW Global Brands, Houston, TX, USA)^[38]^ was used to coat the channel surface for nanoplug assembly/mixing and picodroplet generation.

### Oil combination optimization

We have optimized our oil combination to maximize the retention of resazurin and resorufin molecules within picodroplets, which has been reported to be a common challenge in droplet microfluidic platforms. Towards this end, we examined 3 combinations of oils and surfactants and found that using HFE 7500 oil (3M, St. Paul, MN, USA) as the nanoplug assembly oil and HFE 7500 + 1.2% 008-FluoroSurfactant (RAN Biotechnologies, Inc., Beverly, MA, USA) as the picodroplet generation oil resulted in the most stable generation of picodroplets with the highest retention of resazurin and resorufin (**Table S1**).

### Device Operation

Prior to creating nanoplugs and picodroplets, we filled and primed nanoplug assembly/mixing channel with HFE 7500 oil, which was found to be effective to facilitate nanoplug assembly and mixing. We also filled incubation and detection region of the device with the droplet generation oil (HFE 7500 + 1.2% 008-FluoroSurfactant). Both oils were initially loaded into Tygon microbore tubing (0.02-inch ID and 0.06-inch OD; Cole-Parmer, Vernon Hills, IL, USA) and connected to the device via their designated inlet. We then connected bacteria sample, MH culture broth (with 200 µM resazurin), four antibiotic solutions (including SXT, CIP, GET, TOB, with 200 µM resazurin in each) with corresponding sample and reagent inlets. During device operation, all the inlets on the device were kept under constant pressure, with distinct pre-determined input pressures for the nanoplug assembly oil (3.6 psi), picodroplet generation oil (3.8 psi), samples, MH culture broth and antibiotic solutions (0.9 psi).

To assemble nanoplugs with different antibiotics at multiple dilutions, the integrated microvalves were opened for specific inlets so that the corresponding sample, MH culture broth and/or antibiotic solutions were injected into assembly channel and merged together. By precisely controlling the valve opening time and the ratio between different inputs, the injected antibiotic and its concentration were controlled on demand. The assembled nanoplugs immediately flowed through the serpentine channel to enhance the mixing of different components before picodroplet generation.

To generate picodroplets for single-cell analysis, the mixed nanoplug was pushed into the flow-focusing junction. Then, the pressurized picodroplet generation oil discretized the incoming nanoplug into tens of thousands of picodroplets. Following this step, a PBS barrierplug was introduced at the tale of each droplet group to prevent droplet dispersion and droplet group mixing. The cycle of nanoplug assembly - picodroplet generation - barrierplug introduction was repeated to generate multiple picodroplet groups to test different antibiotics at clinically relevant titrations as well as incorporate spatial coding and no-antibiotic control group, similar to an assembly line. All droplet groups were propelled by the motion of the pressurized picodroplet generation oil and passed through droplet incubation and detection in a continuous flow.

For experiments testing multiple antibiotics in a single device, we added 2 groups of “sacrificial” picodroplets before the 4/6 groups of picodroplets for each antibiotic. These picodroplets, which were excluded from the analysis, were a group of low-fluorescence “spatial coding” picodroplets containing only MH broth to signal the beginning of a new antibiotic and an extra group of no-antibiotic control picodroplets to prevent the high antibiotic titration from the previous antibiotic from interfering with the no-antibiotic control.

### Computational Fluid Dynamics (CFD) of the Flow of a Barrierplug and Droplet in A Microfluidic Channel

Two-dimensional multiphase fluid simulations of a big barrierplug and a picodroplet flowing in a straight microfluidic channel were conducted with a CFD package (COMSOL Multiphysics®, version 5.5, COMSOL Inc.). A laminar two-phase flow, moving mesh model was used to solve for the pressure and velocity field and the location of the interface that defines the barrierplug and droplet in water phase, and the rest of the channel in oil phase. In this model, the channel is 500 μm × 4000 μm (width × length); the barrierplug is created by an ellipse with 1000 μm a-semiaxis by 500 μm b-semiaxis partitioned by the channel; a picodroplet is created by circles with radius of 12.5 μm, and located at the right edge of the channel. Initially the microfluidic channel is filled with oil, with the plug and picodroplet in water phase. The inlet condition of the channel was defined by a fully developed flow with average pressure based on the estimated value from the experiment; The outlet condition was defined with a constant pressure equal to the reference pressure level. The boundary condition of the channel wall was Naiver slip with a 0.5 factor of minimum element length. The fluid-fluid interface was applied at the boundary of the water phase, with no mass flux. The wall-fluid interface is at the contact point between the barrierplug and wall and the picodroplet and the wall, with a contact angle of pi/3. The whole geometry was meshed by free quad elements and the complete mesh consists of 5603 domain elements and 504 boundary elements which gave 29782 degrees of freedom to be solved. The simulation was performed by time-dependent solver with the time stepping method of “BDF”. The global method was unscaled with a tolerance factor of 0.05. The time dependent study was performed with a step size of 0.1 s and a length of 4 s. We set time = 0.5 s as time = 0 s for our screenshot showed in Figure 3B since the driving pressure is applied within the first 0.5 s to keep consistent with our real experiment which has pressure applied from time = 0 s.

### Validation of Barrierplug Separation for Picodroplet Groups

To demonstrate barrierplugs could serve as barriers between picodroplet groups by preventing droplet dispersion, we generated picodroplet groups with different fluorescein concentrations (1.25 nM, 2.5 nM, 5 nM, 10 nM). Barrierplugs were introduced to the tail end of each picodroplet group to separate them apart. We took photos of picodroplet groups when they were flowing through the incubation channel. We also recorded the picodroplet group fluorescence at the detection point using LIF detector. Conversely, in a separated incubation channel design (∼20 min incubation time), we generated 4 picodroplet groups with different fluorescein concentrations (1 nM, 2 nM, 4 nM and 8 nM) but no barrierplugs were injected. The reason we used different incubation channel design is because picodroplets flow would significantly slow down as they gradually dispersed and it would take a very long time before the whole picodroplet group could get to the detection point for the 90-min incubation channel. Similarly, we also imaged those picodroplet groups when they were flowing through the incubation channel and we measured picodroplet fluorescence at the detection point.

### Clinical Sample Collection, Preparation and Validation Using Standard Clinical Diagnostics

Urine specimens representing uncomplicated UTI were obtained and analyzed under an approved institutional review board (IRB) protocol at the Johns Hopkins University School of Medicine (#IRB00189525). For SCALe-AST test, we selected the urine samples with relatively higher bacteria load (>50000 CFU mL^-1^) with *E. coli* as the single dominant pathogen. All the samples were deidentified and stored at 4 °C. To start the test, we firstly filtered 0.5 mL urine through a Primecare membrane (S/G, iPOC^DX^, International Point of Care Inc., Toronto, Canada) with a pore size ranging from 5-35 µm to remove non-bacterial impurities followed by filtration through a second filter with a 0.22 µm pore size (Millex-GV Syringe Filter SLGV004SL, Sigma-Aldrich, Inc., St. Louis, MO, USA). The bacteria were trapped by the second filter and they were flushed out and resuspended in 0.5 mL MH culture broth when we reverted the flow direction. This filtration-based bacteria recovery is similar to a previous report^[39]^ in principle but we have optimized the filter choice to achieve maximum recovery efficiency which was characterized with a spike-in urine sample using droplet digital measurement before and after filtration.^[32]^ The collected bacterial suspension was then measured using Nanodrop to evaluate the concentration. An additional centrifugation step was added to further concentrate 10-20 times for clinical samples that has significantly low concentration (OD600 value below 0.01, bacteria concentration usually between 10^4^ to 10^5^ CFU mL^-1^) before they were loaded into a SCALe-AST device for susceptibility testing test. For the standard clinical diagnostics that were done in the Johns Hopkins Hospital Clinical Microbiology Laboratory, 1-10 µL urine was plated for clinical isolation of uropathogens. The rough uropathogens load was estimated after 18-hour of incubation and then each clinically isolated bacterial specimen was identified using MALDI-TOF mass spectrometry (Bruker Daltonics). The AST was performed using broth microdilution (BD Phoenix), which required up to 48 hours from the time of plating.

### Data Acquisition

Nanoplug assembly, picodroplet generation, incubation and detection were closely monitored using an inverted microscope (IX71; Olympus Corp., Tokyo, Japan) with either a 1.25× magnification objective lens (Olympus PlanAPO N 1.25×/0.04 NA) or a 4× magnification objective lens (Olympus UPlanFl 4×/0.13 NA). The photos for device operation in all figures were taken by a digital single-lens reflex (DSLR) camera (EOS 60D; Canon, Inc., Tokyo, Japan) that was mounted on the microscope. The diameter of picodroplets was calculated by measuring its width in pixels and comparing to the fixed width of the incubation channel (500 μm) using ImageJ.^[40]^

A custom-built laser-induced fluorescence (LIF) system was used to measure picodroplet fluorescence with a 552-nm laser excitation source (OBIS, Coherent, Inc.) and a silicon avalanche photodiode detector (APD) (SPCM-AQRH13, ThorLabs). The laser was operated at 1 mW power and was focused into the detection constriction of the device using a 40× objective (Thorlabs RMS40X-PF, NA 0.75, focal depth approx. 0.6 μm). The picodroplet-emitted fluorescence was continuously acquired by the APD with 0.1 ms sampling time and the collected data was recorded using a LabView program for future analysis.

### Data Analysis

The collected data of picodroplet fluorescence was loaded into MATLAB for data analysis. Firstly, spatial coding groups were identified via the significantly low signal for easy pick up of droplet groups of each individual antibiotic (**Figure S8**). Then, fluorescence intensity of each droplet group was fitted using a Double Gaussian Fitting Model to measure the mean and standard deviation of the negative population. Droplet intensities were corrected by aligning the negative population across different antibiotic concentrations. All droplets with intensities over six times of standard deviation above negative mean were counted as bacteria-containing “positive droplets”. To combine droplet intensity and positive droplet percentage for more accurate MIC/category call, we converted the fluorescence measurement of each positive droplet into an intensity score which was determined by subtracting the background threshold (B) from the measured fluorescence intensity (I), normalizing it with the background threshold, and weighting it with the positive rate of its corresponding droplet group R_pos_ (Equation 1):

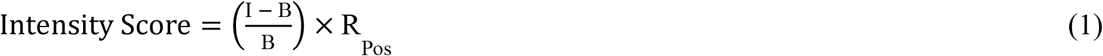

Finally, the intensity score of positive population at each antibiotic concentration was compared with that of no antibiotic control using one-tailed Student’s t-test. With 0.01 as the threshold, we gave the MIC call or category call using the resulted p-value from the Student’s t-test.

## Data Availability

All data is available in Supplementary Material

## Supporting Information

Supporting Information is available from medRxiv.

## Acknowledgements

Research reported in this publication is financially supported by the National Institutes of Health (R01AI137272, R01AI138978, R01AI117032).

Received: ((will be filled in by the editorial staff))

Revised: ((will be filled in by the editorial staff))

Published online: ((will be filled in by the editorial staff))

