## Supplementary Material for "A Cascaded Droplet Microfluidic Platform Enables High-throughput Single Cell Antibiotic Susceptibility Testing at Scale"

**S1. SCALe-AST device design and optimization**

SCALe-AST consists of four different layers, including adaptor layer/cover glass, fluidic layer, valve layer and glass substrate (**Figure 2, Figure S12**). The ‘push-up’ design, namely that the microvalve layer was put under the fluidic layer, was adopted to avoid direct contact between aqueous solution with hydrophilic glass substrate for minimizing nanoplug and picodroplet sticking. To prevent picodroplet evaporation during on-chip incubation, a thin fluidic layer (~1 mm) was assembled where a cover glass was put on top of the incubation channel to fully seal it. As a comparison, when a thick fluidic layer (~5 mm) was used, we observed severe picodroplet evaporation for initial picodroplet groups even though a same sealing cover glass was bonded on top of the incubation channel (**Figure S13**).

Other than the nanoplug mixing channel and flow-focusing junction, the incubation channel height was also optimized to facilitate barrierplug-enabled picodroplet separation. We determined the optimal incubation channel height to be ~60 µm since we found empirically that barrierplug would be prone to stick to channel wall with a lower incubation channel.

**S2. Uniform droplet incubation duration afforded by barrierplugs**

Based on the fluorescence trace of droplet groups with or without barrierplug separation, we measured the time each individual droplet spends in our integrated incubation channel to characterize the uniformity of droplet incubation time. With no barrierplugs, droplet dispersion resulted in a long trail and caused significant broadening of droplet group signal bandwidth (**Figure 3A**). As a result, the coefficient of variation (denoted as CV hereafter, CV = standard deviation / mean) of droplet incubation duration could be as high as 20% even for a short ~20-min incubation channel design (**Figure S6A**). Conversely, in the presence of barrierplugs, droplet groups were tightly packed thus expected to have minimal variations for the droplet incubation duration (**Figure 3C**). Indeed, the CV of droplet incubation duration within one group was below 0.4% (**Figure S6B**) with a maximal 1.5 min difference for a 90-min incubation channel design.

**S3. Time and cost saving using SCALe-AST**

We assume the droplet generation, incubation and detection for testing one antimicrobial condition requires 2 min, 90 min and 2 min, respectively. If the device can only test a single antimicrobial condition per device, we have to start second condition until the first condition test finishes. The total time of testing 4 antibiotics at 6 different titrations is as follows:

$$T_{total}=4*6*\left( T_{Generation}+ T_{Incubation}+T_{Detection} \right)=24*94\min=37.6\mathrm{hours}$$

However, using SCALe-AST, we can immediately start the second condition when the preceding droplet generation is done. The total time of testing same antibiotic/titration conditions will be (here we put 8 droplet groups for each antibiotic since we include an additional spatial coding group and an extra no-antibiotic group):

$$T_{total\_multiplex}=4*8*T_{Generation}+T_{Incubation}+T_{Detection}=\left( 32*2+90+2 \right)min=2.6 hours$$

Therefore, SCALe-AST can significantly shorten the downtime for each antibiotic condition and is capable of testing multiple clinically-relevant antibiotic concentrations in a timely manner. Moreover, SCALe-AST can test tens of different droplet groups in a single device while standard droplet device requires an additional device for each new condition. Thus, SCALe-AST also contributes to cost savings.

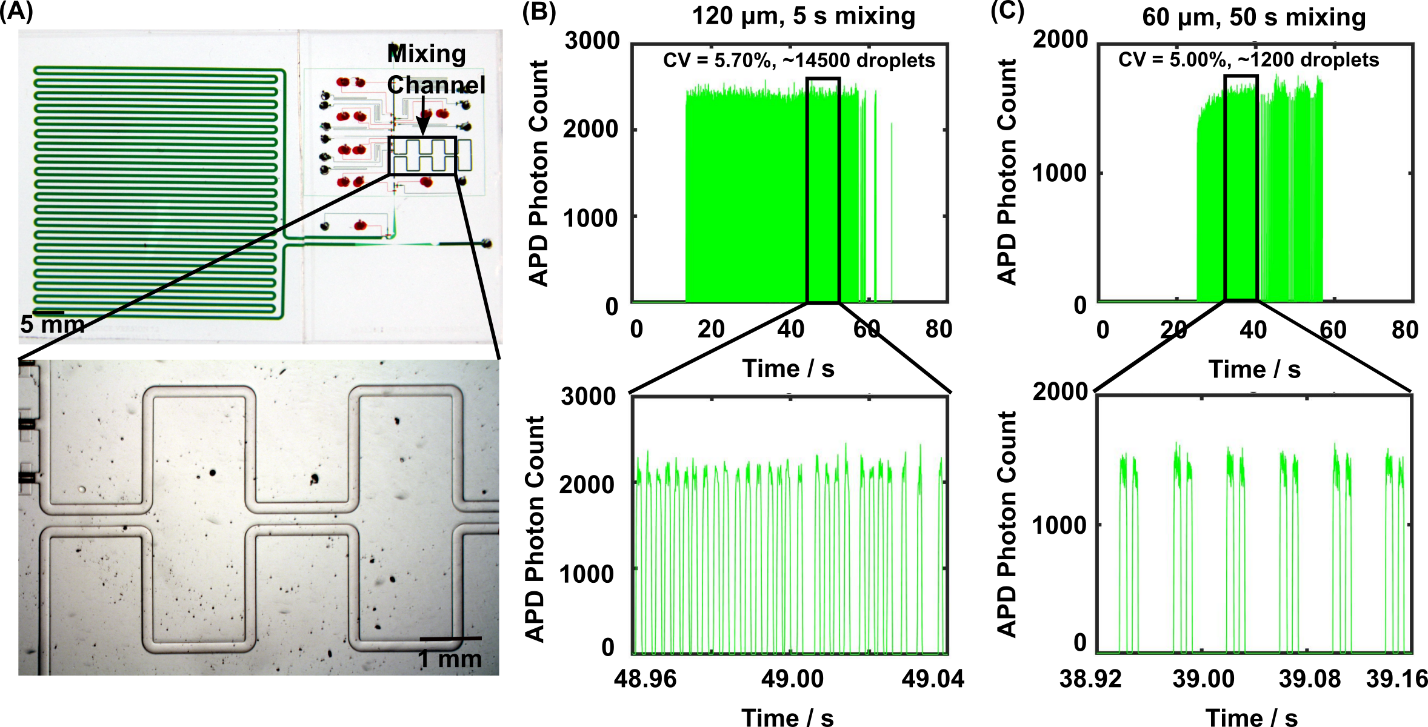

**Figure S1.** **The optimized mixing channel design enables rapid and thorough mixing.**  (A) The mixing channel locates at the immediate downstream of assembly channel. To characterize the nanoplug mixing, we assembled PBS and fluorescein into a nanoplug and discretized it into picodroplets after mixing. We then measured the fluorescence of these picodroplets immediately after generation to characterize the mixing. (B) The fluorescence trace of picodroplets showed uniform intensity (measured using avalanche photodiode (APD) photon count) across ~14000 droplets in a single group after 5 s mixing when the channel height was ~120 µm, indicating the mixing was thorough. This result was similar to the result after ~50 s mixing when a longer but lower mixing channel (60 µm) was used (C).

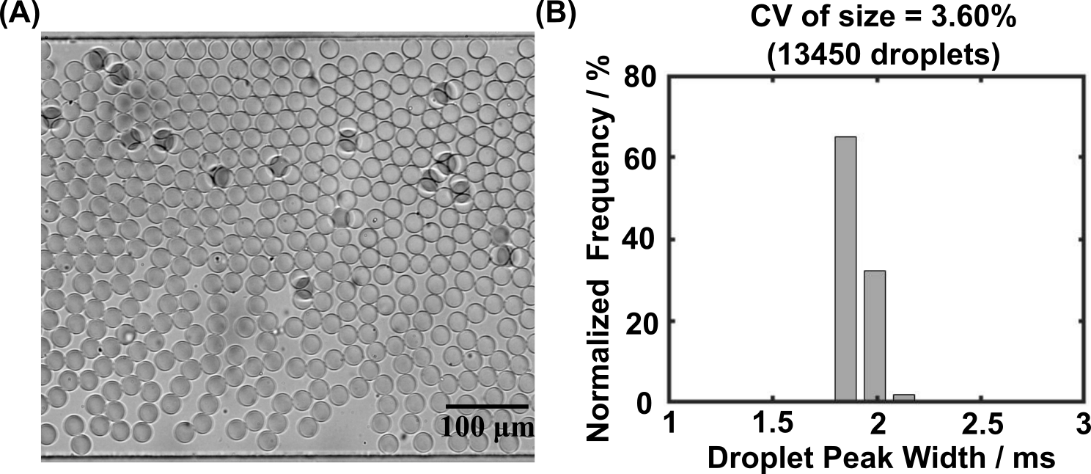

**Figure S2. SCALe-AST generates uniform droplets.** (A) The microscope image shows uniform size of droplets with a volume of ~8 pL. (B) The 3.6% CV (calculated from 13450 picodroplets) demonstrates picodroplets generated in SCALe-AST are uniform.

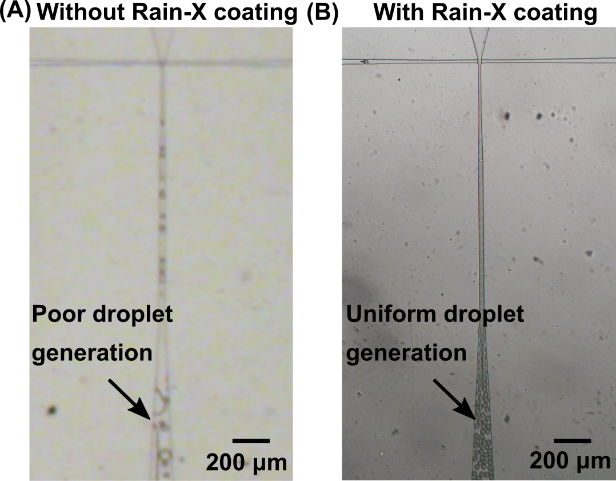

**Figure S3.** **Rain-X coating stabilizes droplet generation**. To guarantee stable picodroplet generation, we found that coating the flow-focusing junction with Rain-X was essential. To show the benefit of Rain-X coating, we used the devices with or without Rain-X coating to generate picodroplets from the assembled nanoplugs. (A) Droplet generation was unstable and the generated droplets were heterogeneous in size without Rain-X modification; (B) Droplet generation was stable after the flow-focusing junction was coated using Rain-X and the generated droplets were uniform.

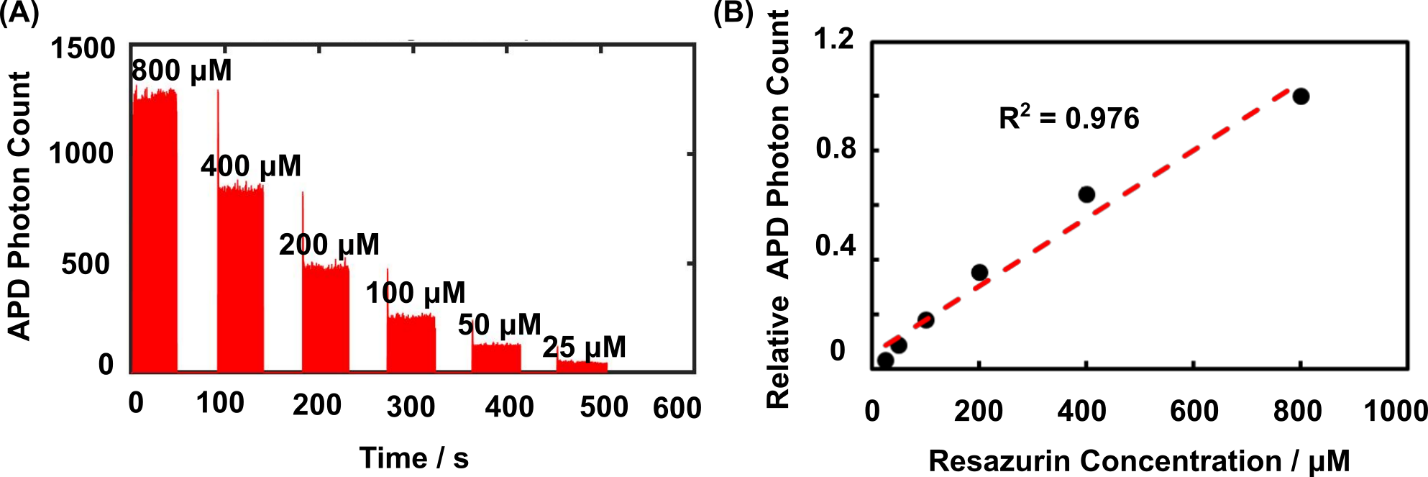

**Figure S4.** **SCALe-AST allows highly-precise titration generation in picodroplet groups.**  To demonstrate this, we assembled Mueller Hinton (MH) broth and resazurin into 6 nanoplugs with different resazurin concentrations and discretized them into picodroplets after mixing. (A) We measured the fluorescence of these picodroplets immediately after generation and the fluorescence trace showed gradually decreased fluorescence with lower resazurin concentrations. The higher peaks in the beginning of each droplet group were the left over from previous group with higher resazurin concentration and only accounted for less than 1% of total droplets, which had negligible effect on our result. (B)The detected fluorescence intensity (normalized APD photon count) increased with the resazurin concentration with a high linearity (n=2, error bar is too small and invisible here), suggesting SCALe-AST could precisely control the content of each nanoplug and thus each picodroplet group.

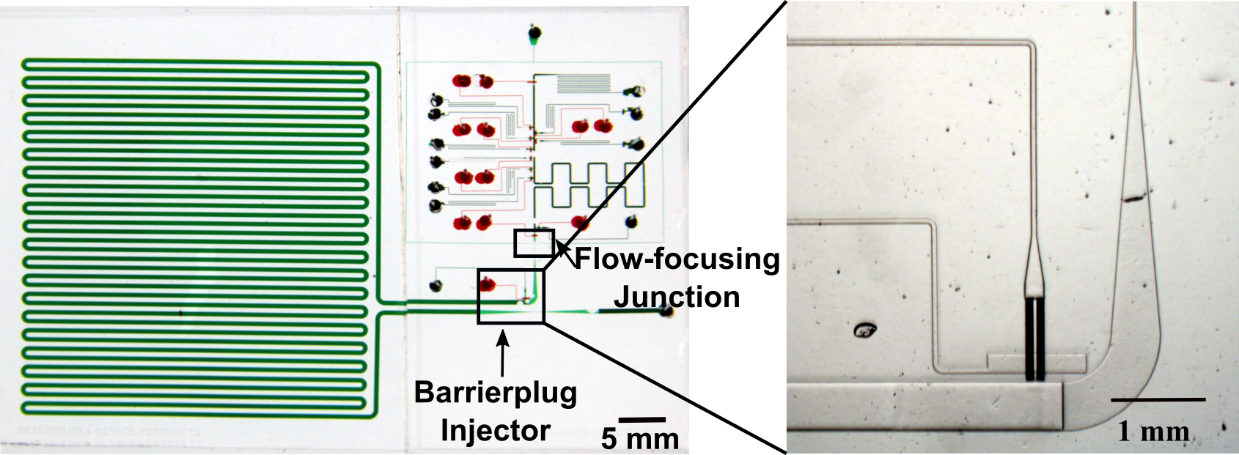

**Figure S5.** **The barrierplug injector introduces barrierplugs into tail end of each picodroplet group via programmable microvalve actuation**. The barrierplug injector is located downstream of the flow-focusing junction. The pressurized PBS can be injected through microvalve actuation and form a barrierplug at the tail end of each picodroplet group to prevent unwanted mixing between picodroplet groups.

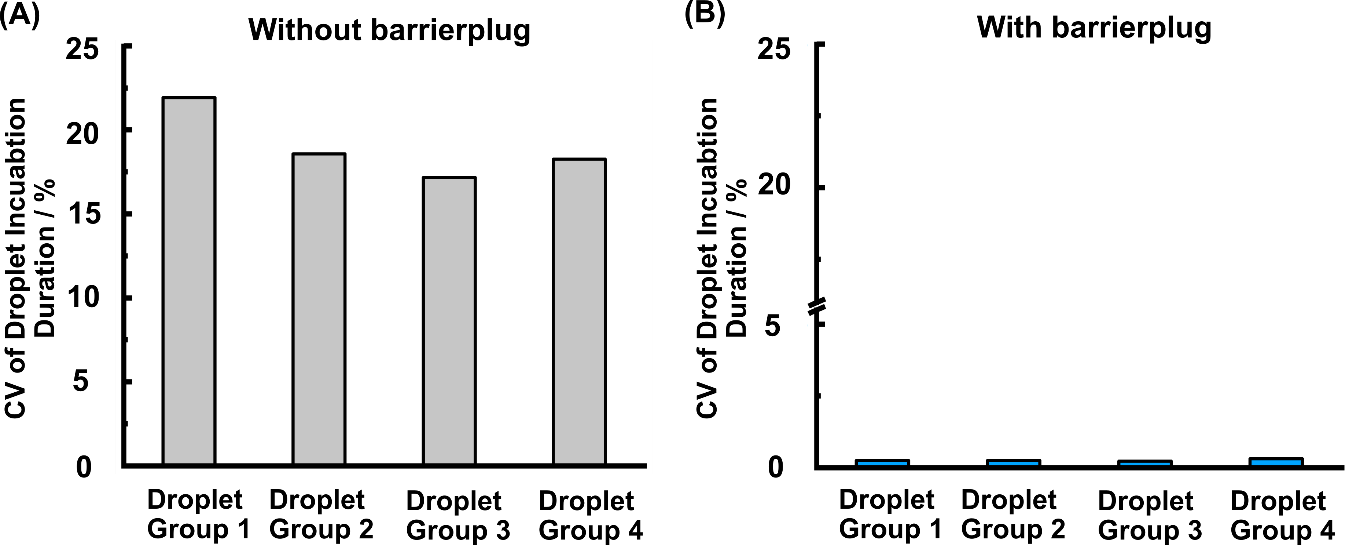

**Figure S6.** **The barrierplugs enables** **uniform incubation duration of droplet groups.** (A)The coefficient of variation (CV) of droplet incubation duration can be as high as 20% for a 20-min incubation channel design in the absence of barrierplugs. (B)In the presence of the barrierplugs, the CV of droplet incubation time within one group drops below 0.4% for a 90-min incubation channel.

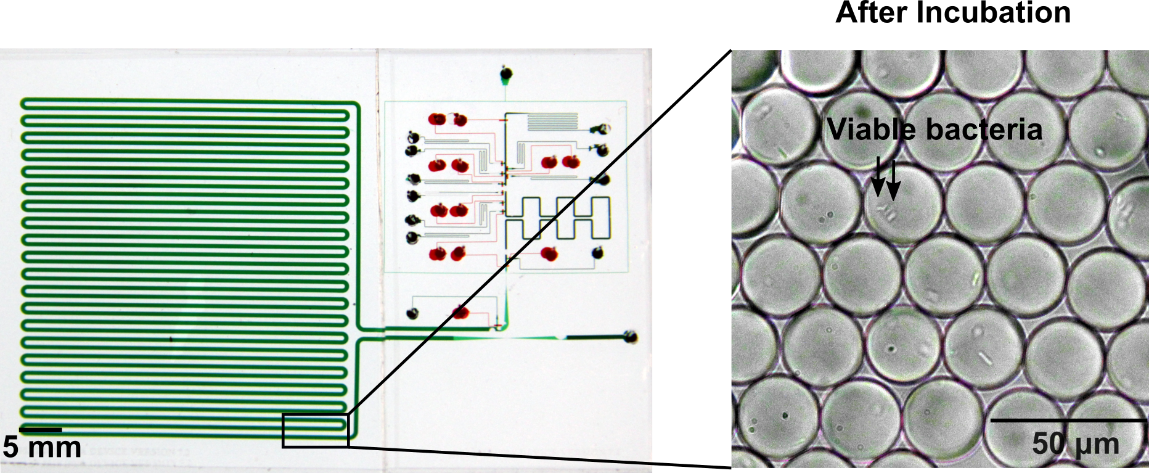

**Figure S7.** ***E. coli* grow in picodroplets**. Viable bacteria were found in the picodroplets at the end of incubation channel when no antibiotic was added. Bacteria typically at least doubled from the digitally encapsulated single bacteria after on-chip 37 ⁰C incubation.

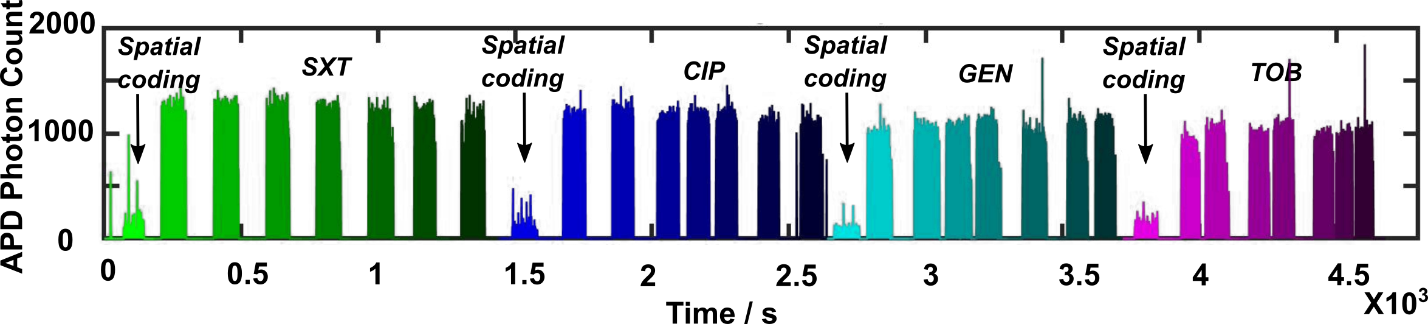

**Figure S8.** **SCALe-AST enables generation of 32 picodroplet groups in a single device for MIC measurement.** Four antibiotics (including trimethoprim-sulfamethoxazole (SXT), ciprofloxacin (CIP), gentamicin (GEN), and tobramycin (TOB)) are co-encapsulated at different concentrations with bacteria, resazurin as well as MH broth. For each antibiotic, we have a spatial coding group containing only MH broth with a low fluorescence signal, two no-antibiotic groups and 5 groups of different antibiotic concentrations that all contain resazurin and present high fluorescence signals. In total, 32 (4×8) droplet groups are generated in a single SCALe-AST device.

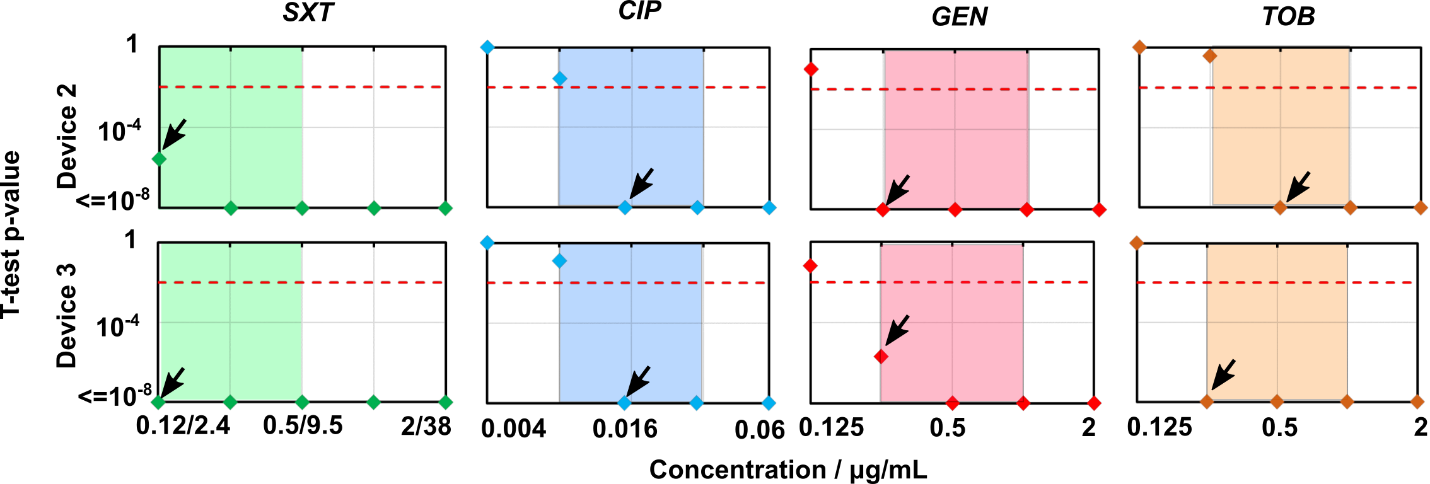

**Figure S9.** **SCALe-AST shows consistent MIC measurements for the ATCC reference *E. coli*.** Using p-value derived from t-test, our SCALe-AST identifies MICs for the ATCC reference *E. coli* against four common antibiotics (including SXT, CIP, GEN and TOB) in ~2.5 hours in two additional independent devices and the results remain consistent with CLSI MICs.

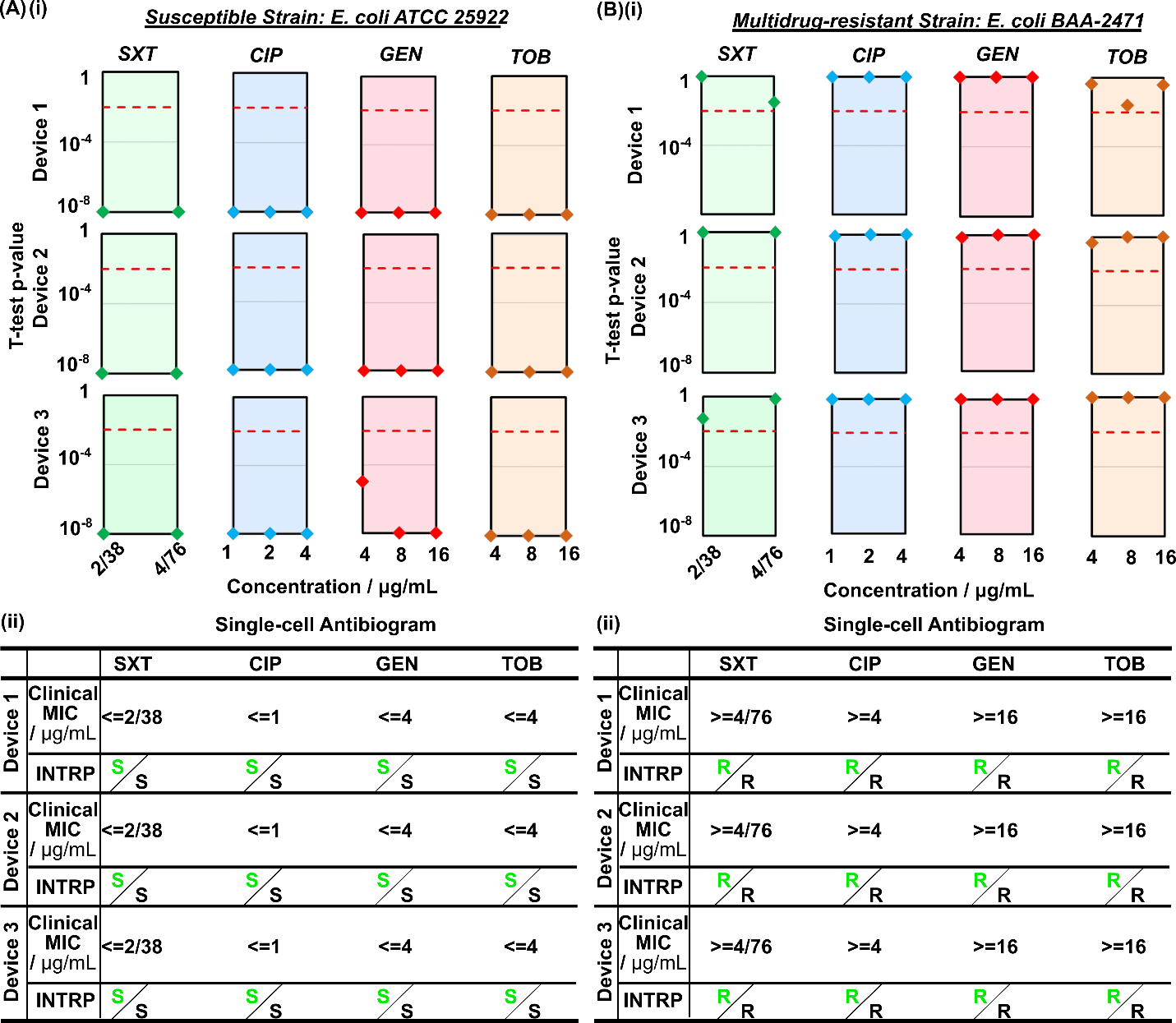

**Figure S10.** **Interpretive breakpoint testing reliably classifies reference *E. coli* strains**. (A)(i) The t-test p-values for interpretive breakpoint testing using susceptible reference strain all fell below the threshold and showed a significant difference between antibiotic treatment and no antibiotic control, indicating bacterial growth was significantly inhibited. Therefore, the clinical MICs for SXT, CIP, GEN and TOB were identified as lower than or equal to 2/38 µg mL^-1^, 1 µg mL^-1^, 4 µg mL^-1^, and 4 µg mL^-1^, respectively, indicating the bacteria were susceptible to all four antibiotics based on CLSI guideline, matching with susceptible phenotype (ii). The black S indicates the susceptible phenotype of *E. coli* reference strain while the green S means agreement of SCALe-AST category with bacterial phenotype. (B)(i) The t-test p-value of interpretive breakpoint testing using a multidrug-resistant strain were all above the threshold and showed no significant difference between antibiotic treatment and no antibiotic control, suggesting the bacteria growth was negligibly affected. Thus, the measured clinical MICs were above 4/76 µg mL^-1^, 4 µg mL^-1^, 16 µg mL^-1^, and 16 µg mL^-1^ for SXT, CIP, GEN and TOB, respectively, suggesting the bacteria were resistant to all four antibiotics, which again was consistent with the multidrug-resistant phenotype (ii). The interpretive breakpoint tests for both susceptible and multi-drug resistant reference strains were tested in triplicate in different devices to demonstrate the robustness of SCALe-AST.

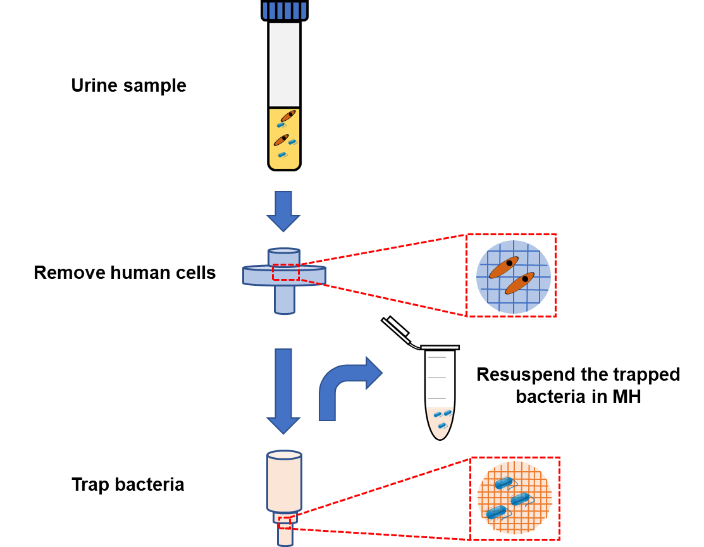

**Figure S11.** **The facile filtration protocol facilitates rapid bacteria recovery from urine specimens.** The urine sample is firstly filtered to remove large non-bacterial impurities using a filter with a pore size ranging from 5 µm to 35 µm followed by a second filtration to trap the bacteria on the filtering membrane with a 0.22 µm pore size. The flow direction is then reverted to resuspend the trapped bacteria in MH broth for interpretive breakpoint testing.

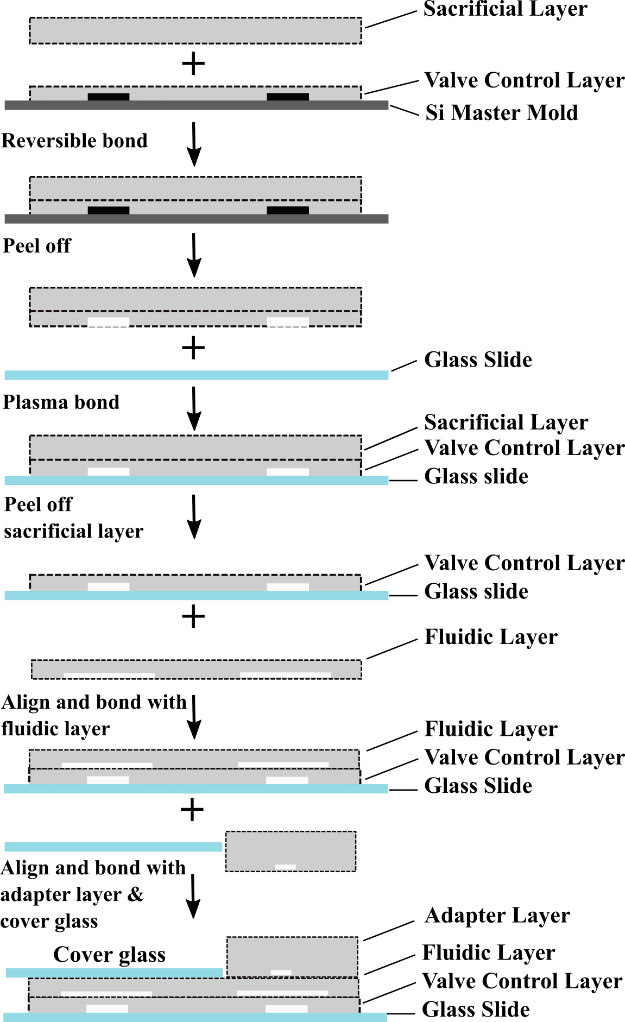

**Figure S12.** **The SCALe-AST fabrication protocol.** A thin valve layer was fabricated and peeled from the mold with the help of a reversibly-bonded thick sacrificial layer followed by bonding with glass slides. The sacrificial layer was discarded before bonding a fluidic layer on top of the thin valve layer. Finally, an adapter layer was stacked on top of microvalve part of fluidic layer while the incubation channel was sealed with a cover glass.

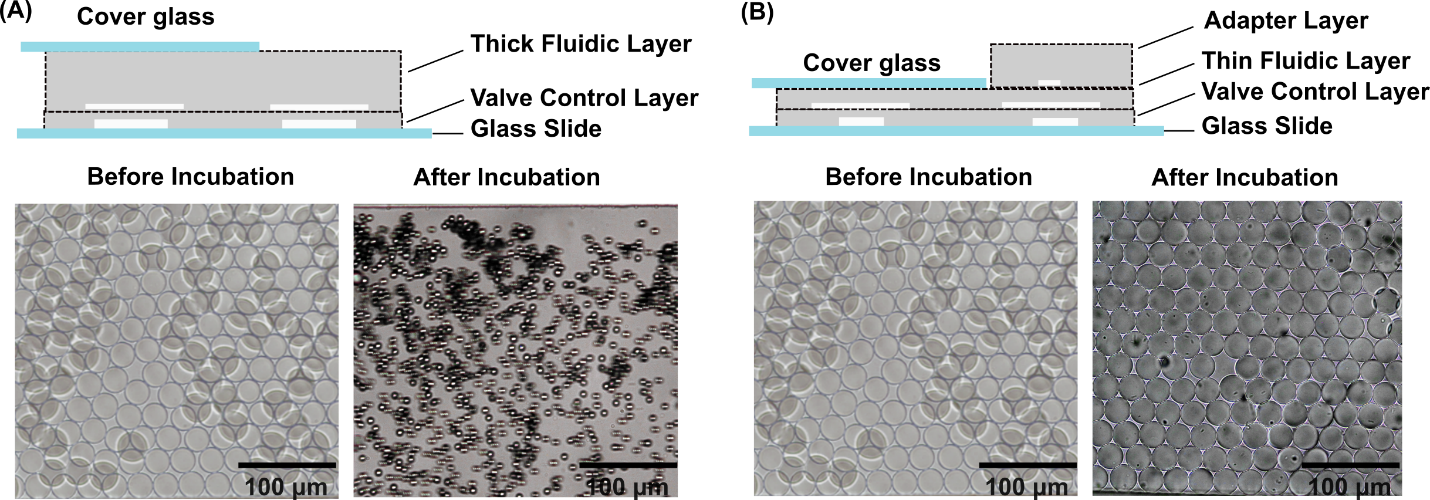

**Figure S13.** **The thin fluidic layer sealed by a cover glass prevents picodroplet evaporation.** (A) The thick fluidic layer has a larger side interface which results in a severe evaporation for initial droplet groups after 90 min incubation at 37 ⁰C. (B) By using a thin fluidic layer instead sealed with a cover glass, the side interface is minimized and as a result, the evaporation is prohibited.

**Table S1. Optimization of oil combination for SCALe-AST**

Using HFE 7500 for nanoplug assembly and HFE 7500 plus 1.2% 008-FluoroSurfactant for droplet generation ensure stable droplet generation as well as minimal resazurin diffusion. Other combination would result in either unstable droplets or fluorophore diffusion.

| **Nanoplug assembly oil** | **Droplet generation oil** | **Nanoplug Assembly** | **Picodroplet Outcome** |
| --- | --- | --- | --- |
| FC40 | BioRad Droplet Generation Oil | Stable nanoplugs | Fluorophore diffusion |
| FC40 | HFE 7500 + 1.2% 008-FluoroSurfactant | Stable nanoplugs | Unstable droplets |
| HFE 7500 | HFE 7500 + 1.2% 008-FluoroSurfactant | Stable nanoplugs | Stable droplets & Minimal diffusion |
